# MRI-derived brain aging trajectories across the Alzheimer’s continuum: links to cognition, pathology and lifestyle risk factors

**DOI:** 10.64898/2026.08.19.26360525

**Authors:** Elizabeth Kuhn, Georgios Antonopoulos, Luca Kleineidam, Melina Stark, Sandra Roeske, Felix Hoffstaedter, Laura Waite, Oliver Peters, Julian Hellmann-Regen, Lukas Preis, Daria Gref, Josef Priller, Eike Jakob Spruth, Maria Gemenetzi, Anja Schneider, Klaus Fliessbach, Jens Wiltfang, Björn H. Schott, Franziska Maier, Emrah Düzel, Wenzel Glanz, Enise Incesoy, Renat Yakupov, Falk Lüsebrink, Katharina Buerger, Daniel Janowitz, Sophia Stöcklein, Robert Perneczky, Boris-Stephan Rauchmann, Stefan J. Teipel, Ingo Kilimann, Christoph Laske, Sebastian Sodenkamp, Annika Spottke, Frederic Brosseron, Aflredo Ramirez, Matthias C. Schmid, Stefen Hetzer, Peter Dechent, Frank Jessen, Simon B. Eickhoff, Kaustubh R. Patil, Michael Wagner, the Alzheimer’s Disease Neuroimaging Initiative, the DELCODE study group

## Abstract

**Background:** The brain age gap (BAG), the difference between neuroimaging-predicted and chronological age, captures inter-individual variation in brain aging. Although sensitive to Alzheimer’s disease (AD) pathology, its longitudinal patterns across the clinical AD continuum and prognostic relevance remain unclear.

**Methods:** 577 participants from the DELCODE cohort (>2,100 MRI scans) were analysed: healthy controls individuals (HC, N=202), and patients with subjective cognitive decline (SCD, N=248), mild cognitive impairment (N=93), and AD dementia (N=34). All underwent structural MRI, amyloid (Aβ_42/40_) and phosphorylated tau181 assessment, and lifestyle-related dementia risk profiling (LIBRA). BAG was derived using brainageR. Associations with baseline cognition, cognitive decline, and clinical progression (up to eight years) were examined using mixed-effects and Cox models. Mediation analyses tested whether BAG accounted for LIBRA-cognition associations. Biomarker-related and clinical findings were replicated in ADNI (N=461).

**Findings:** BAG showed excellent short-term reliability, increased stepwise across the clinical spectrum and was elevated in amyloid-positive SCD, but not in asymptomatic amyloid-positive HC. Longitudinal BAG increases were strongest in amyloid- and tau-positive participants (Aβ+T+). Higher BAG was associated with poorer baseline cognition and predicted cognitive decline, with strongest effects in Aβ+T+. All main findings replicated in ADNI. BAG was associated with LIBRA only in biomarker-negative participants and partly mediated associations with cognitive outcomes in DELCODE.

**Interpretation:** BAG is a reliable non-invasive marker of structural brain health sensitive to AD pathology and to modifiable AD risk. Detectable divergence prior to objective cognitive impairment supports its relevance for early risk stratification and prevention-oriented research.

**Funding:** Helmholtz AI Cooperation Unit (ZT-I-PF-5-163).

## Introduction

Aging is the strongest risk factor for neurodegenerative diseases, including Alzheimer’s disease (AD). Structural and functional brain changes occurs during normal aging, but are also characteristic of neurodegenerative conditions, making it challenging to distinguish healthy from pathological brain aging.^1^ This overlap has led to the view that neurodegenerative diseases represent manifestations of accelerated brain aging, although accumulating evidence also supports the contribution of disease-specific biological processes to their onset and progression.^2,3^ Brain aging shows marked inter-individual variability: individuals of the same chronological age can exhibit substantially different brain phenotypes, shaped by genetics background, lifestyle exposures, and chronic health conditions.^4,5^ Advances in machine learning applied to neuroimaging now allow estimation of an individual’s “brain age,” from which the Brain Age Gap (BAG), the difference between predicted and chronological age, can be derived.^6,7^ A higher BAG reflects accelerated brain aging and has been linked to increased mortality, neurodegenerative and psychiatric disorders, and other clinical conditions.^8–10^

In AD, elevated BAG has been observed from the mild cognitive impairment (MCI) stage and linked to increased risk of progression from MCI to major neurocognitive disorder (MND).^9,11,12^ However, most evidence remains cross-sectional and confined to specific clinical stages. Although some studies have examined associations with longitudinal outcomes, BAG trajectories across the full AD continuum, particularly at the earliest, preclinical stages, are largely unknown. Findings in cognitively unimpaired (CU) older adults are inconsistent regarding association with AD biomarkers,^13–15^ and patients with subjective cognitive decline (SCD) recruited from memory clinics have not been specifically examined.

Moreover, whether within-person changes in BAG over time reflect ongoing neurodegenerative processes, particularly in the presence of underlying amyloid and tau pathology (A/T profiles) remains unclear. Clarifying the temporal dynamics of BAG is therefore essential to determine its potential relevance for prognosis and clinical monitoring.

In this study, we therefore characterized BAG across the clinical-biological continuum of AD^16^ and examined its longitudinal trajectories. Using repeated MRI and biomarker assessments, we investigated (i) BAG increases across clinical stages and A/T profiles, (ii) its associations with cognition, cognitive decline, and clinical progression beyond established AD risk markers, and (iii) its relationships with lifestyle-related dementia risk (operationalized by the Lifestyle for BRAin Health, LIBRA score)^17^. By integrating clinical, biomarker, and lifestyle dimensions, this study evaluates whether BAG provides incremental and longitudinally meaningful information relevant for early risk stratification and disease monitoring.

## Methods

### Study design and participants

We analysed data from 577 older adults enrolled in the *Longitudinal Cognitive Impairment and Dementia Study (DELCODE)*, a prospective, multicentre cohort coordinated by the German Center for Neurodegenerative Diseases (DZNE).^18^ Recruitment occurred across ten university-based memory clinics in Germany. DELCODE is registered in the German Clinical Trials Register (DRKS00007966), approved by local ethics committees, and all participants provided written informed consent. This report follows *STROBE* guidelines for observational cohort studies.

Participants were either community volunteers (healthy controls [HC], n=202) or patients referred to memory clinics with subjective cognitive decline (SCD, n=248), mild cognitive impairment (MCI, n=93), or mild Alzheimer-type MND (DAT, n=34). Diagnostic classification was harmonized across sites using the *Consortium to Establish a Registry for Alzheimer’s Disease (CERAD)* battery for ensuring absence of cognitive impairment (in SCD and HC) or presence of amnestic MCI. SCD was defined by self-reported memory concerns with unimpaired neuropsychological performance (>−1·5 SD). MCI required impaired episodic memory (<−1.5 SD on the CERAD word list delayed recall trial) consistent with amnestic MCI criteria. Mild DAT was diagnosed clinically with Mini-Mental State Examination (MMSE) ≥18.

Inclusion criteria were age ≥60 years, German fluency, and availability of a study partner. Exclusion criteria were major psychiatric disorders, non-AD neurodegenerative diseases, stroke with residual symptoms, unstable medical disease, active malignancy, or chronic psychoactive or antidementia medication use. For the present analyses, participants also required baseline data for: (1) structural MRI, (2) fluid biomarkers (cerebrospinal fluid [CSF] or plasma Aβ_42/40_ ratio and phosphorylated tau181 [ptau_181_]), (3) LIBRA lifestyle risk scores, and (4) longitudinal follow-up enabling clinical progression assessment in non-demented participants (details in **appendix p2)**.

### Magnetic resonance imaging and brain age estimation

MRI was performed at nine sites on Siemens’s scanners (3 TIM Trio, 4 Verio, 1 Skyra, 1 Prisma). High-resolution T1-weighted structural images (MP-RAGE, TR 2200 ms, TE 2 ms, matrix 256×256, voxel size 1×1×1 mm^3^, flip angle 8°) were acquired with harmonised protocols and underwent visual quality control. Longitudinal MRI (up to four scans per participant, acquired at approximately annual intervals) were available for a subset of 540 [93·6%] participants (median follow-up 3·8 years [IQR 2·2-4·0]; range 0·9-4·6), enabling assessment of within-person trajectories of brain ageing.

Brain-predicted age was estimated using brainageR v2.1 (https://github.com/james-cole/brainageR; containerized version https://github.com/fprados/brainageR_dockerfile), combining voxel-based segmentation and spatial normalisation (SPM12) with Gaussian process regression trained on over 3000 healthy individuals. To minimise systematic bias between predicted and chronological age, we applied the statistical bias correction described by *de Lange et al.* (2019), based on the linear relationship between predicted and chronological age in the control group.^19^ BAG was calculated as MRI-predicted and then bias corrected age *minus* chronological age; positive values indicate an older-appearing brain.

Model performance in HC participants yielded a MAE of 7·5 years and RMSE of 9·2 years (r=0·50), consistent with published performance of brainageR in older adults, clinically enriched cohorts. Calibration of MRI-derived predicted age against chronological age yielded a MAE of 3·6 years and RMSE of 4·5 years (r=0·55; **appendix p4**). In the dedicated test-retest subset (n=130; interval ≤45 days), BAG demonstrated excellent short-interval reliability (ICC(2,1)=0·98, 95% CI 0·97-0·98; r=0·98).

### Cognition and clinical progression

Global cognition was assessed with the Preclinical Alzheimer’s Cognitive Composite (PACC5),^20^ defined as the mean of standardized z-scores from: Free and Cued Selective Reminding Test (FCSRT; total and free recall, 0-96), Symbol Digit Modalities Test (SDMT, 0-90), Logical Memory delayed recall (0-25), semantic fluency (animals + groceries in 1 min, 0-60), and the MMSE (0-30). Higher scores reflect better performance.

Incident MCI was identified using a two-step process adapted from the Wisconsin Registry for Alzheimer’s Prevention, combining algorithmic screening with expert review by neuropsychologists blinded for group assignment (FCSRT total ≤46, MMSE ≤26, clinician/informant reports using standard MCI criteria).^21^ Incident MND was determined by a central diagnostic committee integrating cognitive, functional, and clinical information (CDR≥1 or MMSE≤23, study physician confirmation when needed).^22^ Over a median follow-up of 5·0 years (IQR 4·0-6·4, range 0·9-8·3), 23 HC (11·4%) and 65 SCD (26·2%) participants developed MCI (7 subsequently progressed to MND), and 35 MCI participants (37·6%) progressed to MND. Mean time to progression was 4·6 [2·2] years overall.

### Alzheimer’s disease biomarkers

Biomarker classification followed an Aβ/T framework,^23^ using CSF when available and plasma otherwise. Overall, 339 (58·8%) participants had CSF assays and 238 (41·2%) had plasma assays. Cut-offs were derived via Gaussian mixture modelling for CSF (Aβ_42/40_ ≤0·08; p-tau_181_≥73·65 pg/ml). For plasma, the Aβ_42/40_ threshold was obtained by regression-based projection of the CSF cut-off (≤0·106), and the p-tau_181_ threshold by ROC analysis against CSF (≥1·707 pg/mL). Participants were categorised as Aβ-T− (n=327, 56·7%), Aβ-T+ (n=28, 4·9%), Aβ+T− (n=138, 23·9%), or Aβ+T+ (n=84, 14·6%), with tau status defined using core 1 tau (**appendix p2**).

### Lifestyle for brain health (LIBRA)

The LIBRA score quantifies modifiable dementia risk across 12 behavioural and clinical factors: smoking, physical inactivity, obesity, hypertension, hypercholesterolaemia, diabetes, coronary heart disease, depression, renal dysfunction, diet, alcohol consumption, and cognitive activity. Each factor is weighted by its relative contribution to dementia risk, producing a composite score ranging from negative (favourable) to positive (unfavourable) values.^17^

### ADNI replication cohort

Alzheimer’s Disease Neuroimaging Initiative (ADNI) was used as an independent replication cohort; lifestyle analyses were omitted as data required for calculating LIBRA are not available. Participants included 262 HC, 148 MCI (early- and late-MCI merged), and 51 DAT. Amyloid status was defined using centiloid-transformed florbetapir- (N=187 [40·6%]) or florbetaben-PET (N=272 [59·4%]; positivity thresholds 22·5 and 20·2, respectively, see adni.loni.usc.edu pipeline), and tau status using flortaucipir-PET entorhinal SUVr (threshold 1·2; details in **appendix pp2-3**).^24^ ADNI was launched in 2003 as a public-private partnership led by Principal Investigator Michael W. Weiner, MD. The primary goal of ADNI has been to test whether serial magnetic resonance imaging (MRI), positron emission tomography (PET), other biological markers, and clinical and neuropsychological assessment can be combined to measure the progression of mild cognitive impairment (MCI) and early Alzheimer’s disease (AD).

### Statistical analysis

Linear mixed-effects models (lme4, R, version 4.2.3) were used to investigate baseline differences and longitudinal change in BAG across clinical groups (HC, SCD, MCI, DAT), amyloid and A/T profiles, and its association with cognitive performance (PACC5), and lifestyle-related dementia risk (LIBRA). Models included random intercepts for participants and time from baseline, and fixed effects for age, sex, and education, as well as their interactions with time. Group-specific effects and simple slopes were estimated using *ggeffects*, and post-hoc contrasts were corrected for multiple comparisons. BAG-cognition analyses additionally included time x BAG x biomarker-positivity interactions to test whether BAG predicted differential cognitive decline across biomarker-defined subgroups.

Associations between baseline BAG and time-to-event outcomes (progression to incident MCI and/or MND) were modelled using Cox proportional hazards models, with hazard ratios (HR) and 95% confidence intervals reported. Joint longitudinal-survival models were used to confirm robustness to selective attrition and the longitudinal evolution of BAG.

Mediation analyses were performed using structural equation modelling (lavaan, R), testing whether BAG partially mediated the association between LIBRA (predictor) and PACC5 (outcome, at baseline and over time through slopes). Indirect effects were estimated using 1000 bootstrap resamples to obtain bias-corrected confidence intervals. Moderation by biomarker status was evaluated by repeating analyses within Aβ- and Aβ+ subsamples. All analyses were two-sided with significance defined at α=0·05. Sensitivity analyses included adjustment for hippocampal volume and baseline cognitive performances, and restriction to CU participants (HC and SCD).

## Results

We included 577 older adults (mean age 70·8[6·0] years, 292 [50·6%] women): 202 HC, 248 SCD, 93 MCI, and 34 DAT. Baseline demographics, clinical, and biomarker characteristics are shown in **Table 1**.

**Table 1.** Baseline participants demographics according to clinical groups.

|  | HC | SCD | MCI | DAT | Overall | P |
| --- | --- | --- | --- | --- | --- | --- |
| No. (%) | 202 (35.0%) | 248 (43.0%) | 93 (16.1%) | 34 (5.9%) | 577 (100%) |  |
| Age, y | 67.6 (64.7, 72.1) | 70.6 (66.8, 74.9) | 73.8 (69.2, 77.9) | 74.7 (71.0, 78.7) | 70.4 (66.0, 75.0) | <.001 |
| Female | 119 (58.9%) | 114 (46.0%) | 40 (43.0%) | 19 (55.9%) | 292 (50.6%) | .02 |
| Education, y | 14 (12-17) | 15 (13-17) | 13 (12-17) | 12 (11-13) | 14 (12-17) | <.001 |
| MMSE score | 30 (29-30) | 29 (29-30) | 28 (27-29) | 23 (21-27) | 29 (29-30) | <.001 |
| Aβ positivity (%) | 47 (23.3%) | 92 (37.1%) | 54 (58.1%) | 29 (85.3%) | 222 (38.5%) | <.001 |
| A/T positivity (%) | 6 (3.0%) | 32 (12.9%) | 25 (26.9%) | 21 (61.8%) | 84 (14.6%) | <.001 |
| CSF modality (%) | 105 (52.0%) | 145 (58.5%) | 65 (69.9%) | 24 (70.6%) | 339 (58.8%) | .01 |
| FU time, y | 6.1 (4.4, 7.0) | 4.9 (3.9, 6.0) | 4.2 (3.0, 6.1) | 2.1 (1.0, 3.8) | 5.0 (3.4, 6.3) | <.001 |
| FU time MRI, y | 3.9 (3.0, 4.0) | 4.0 (2.1, 4.0) | 3.0 (1.0, 4.0) | 1.1 (1.0, 2.3) | 3.4 (2.0, 4.0) | <.001 |

Baseline values are median (IQR) for continuous data and number of observations (%) for categorical data. *Abbreviations:* Ab, amyloid; FU, Follow-up, IQR, interquartile;
MMSE, mini mental state examination; A/T, amyloid and tau status combined profiles.

### BAG across clinical and biomarker groups

At baseline, BAG differed significantly across clinical groups (overall p<0·0001), with higher values in SCD, MCI, and DAT relative to HC. The largest difference was observed in DAT (approximately +9·6 [5·4 to 13·8] years vs HC), with a stepwise increase along the clinical continuum (**Figure 1A**). When considering amyloid status (i.e., clinical-biological AD construct), BAG was not elevated in Aβ+HC compared to Aβ-HC but was significantly higher in all other amyloid-positive symptomatic groups, reaching its maximum in Aβ+DAT (**Figure 1C**). Overall, amyloid positivity was associated with a modest but significant increase in BAG (+1·5 [0·0-3·0] years; p=0·04), and a similar gradient was observed when combining amyloid and tau status (+2·6 [0·0-5·2] years vs Aβ-T−; p=0·05; **Figure 1E**).

**Figure 1.**
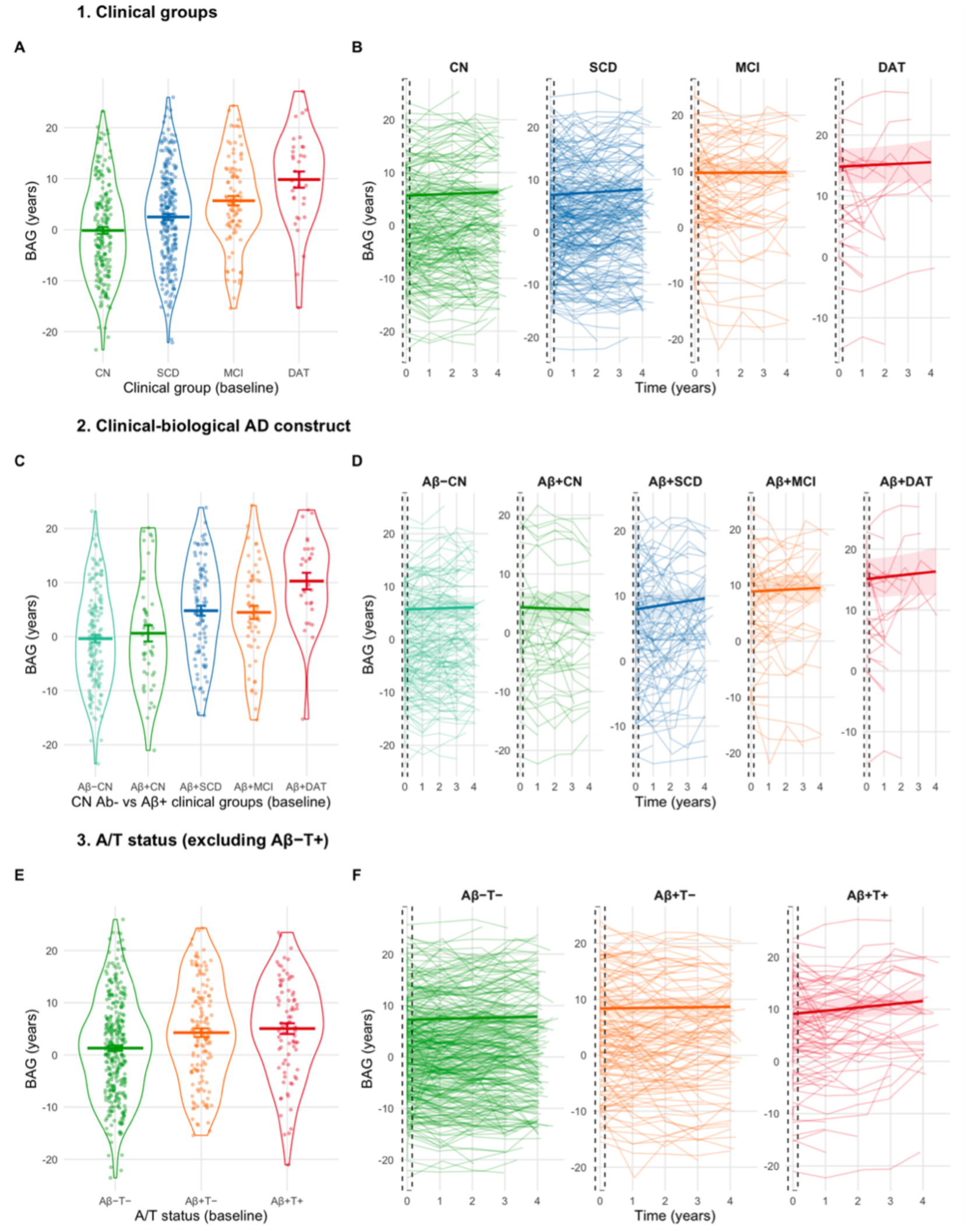
Baseline and longitudinal patterns of Brain Age Gap (BAG) across clinical, amyloid, and A/T groups. Notes. Left panels show baseline BAG distribution across clinical groups (A), clinical-biological AD subgroups (C), and A/T profiles (E). Right panels (B, D, F) display corresponding individual longitudinal BAG trajectories; the dashed box indicates the baseline window (t=0). Across groups, BAG increased stepwise with clinical severity, was higher in amyloid-positive symptomatic groups, and reached its highest values in Aβ+T+ individuals. Abbreviations: HC=healthy controls, SCD=subjective cognitive decline, MCI=mild cognitive impairment, DAT=Alzheimer’s type dementia, BAG=brain-age gap, Aβ=β-amyloid, T=tau.

Across all participants, BAG increased modestly over time (+0·1 [0·0-0·2] years per year; p=0·02). Slopes did not differ across clinical groups nor between amyloid-positive and amyloid-negative participants but accelerated significantly in Aβ+T+ participants (+0·6 [0·1-1·0] years per year vs Aβ-T−; and +0·6 [0·2-1·1] vs Aβ+T−; both p=0·003; **Figure 1 [right panels]**). The same pattern of accelerated BAG trajectories in Aβ+T+ participants was also observed in the CU subsample at a trend level (**appendix pp5–7**).

### BAG and cognition

Higher BAG at baseline was associated with lower PACC5 performance at baseline, with each additional BAG year corresponding to lower scores (β=−0·04, 95% CI −0·05 to −0·03 points; p<0·0001). These associations were stronger in Aβ+ participants (additional BAG-PACC5 slope difference β −0·02, −0·04 to −0·002; p=0·03), and most pronounced in Aβ+T+ individuals (β −0·04, −0·08 to 0·001 vs Aβ+T−; p=0·03).

Higher BAG at baseline predicted faster PACC5 decline (BAG×time interaction, β −0·006; 95% CI −0·008 to − 0·004; p<0·0001), with stronger effects in the Aβ+ group (β −0·005;-0·009 to −0·002, p=0·005). The association was further strengthened in the Aβ+T+ compared with the Aβ+T− group (three-way interaction p=0·005), with the between-group difference increasing from 0·14 (95% CI 0·02–0·30; p=0·008) at low BAG to 0·24 (0·11– 0·37; p<0·001) at high BAG. Findings were unchanged after adjustment for hippocampal volume (**Figure 2**), and were largely replicated in the CU subsample (**appendix pp8-9).**

**Figure 2.**
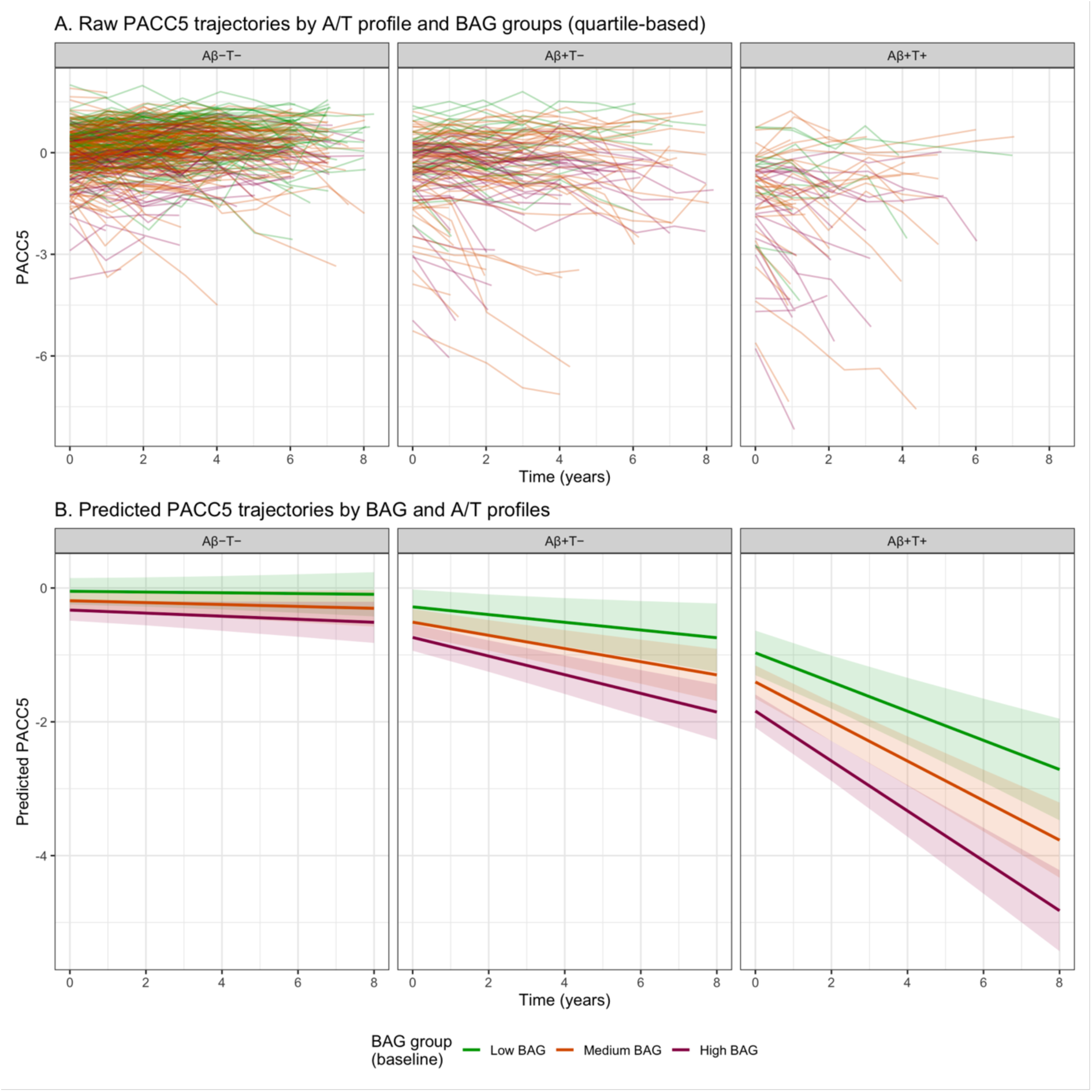
Longitudinal PACC5 trajectories according to brain-age gap (BAG) and A/T profile. Notes. Panel A shows observed individual PACC5 trajectories over follow-up, stratified by A/T profile (Aβ−T−, Aβ+T−, Aβ+T+) and baseline brain-age gap (BAG) groups defined by quartiles (low, medium, high). Panel B shows predicted PACC5 trajectories from linear mixed-effects models including interactions between time, continuous BAG, and A/T profile, adjusted for age, sex, and years of education. Shaded areas represent 95% CIs. Higher BAG indicates an older-appearing brain relative to chronological age. Aβ=β-amyloid. T=tau. BAG=brain-age gap.

### BAG and clinical progression

Baseline BAG was associated with clinical progression. Each additional year of BAG corresponded to a 5% higher risk of progression to MCI or MND (HR 1·05, 95% CI 1·03-1·08; **Figure 3**), a 5% higher risk of incident MCI (HR 1·05, 1·02-1·08), and a 9% higher risk of MND (HR 1·09, 1·04-1·13; all p<0·001). There was no evidence of interaction between BAG and amyloid status or A/T profiles (all interaction p>0·05). Associations were attenuated after adjustment for baseline pacc5 scores (HR 1·03, 95% CI 1·00-1·05; p=0·02) and hippocampal volume but remained statistically significant or of similar magnitude (HR 1·02, 95% CI 1·00-1·05; p=0·06; **appendix p10**).

**Figure 3.**
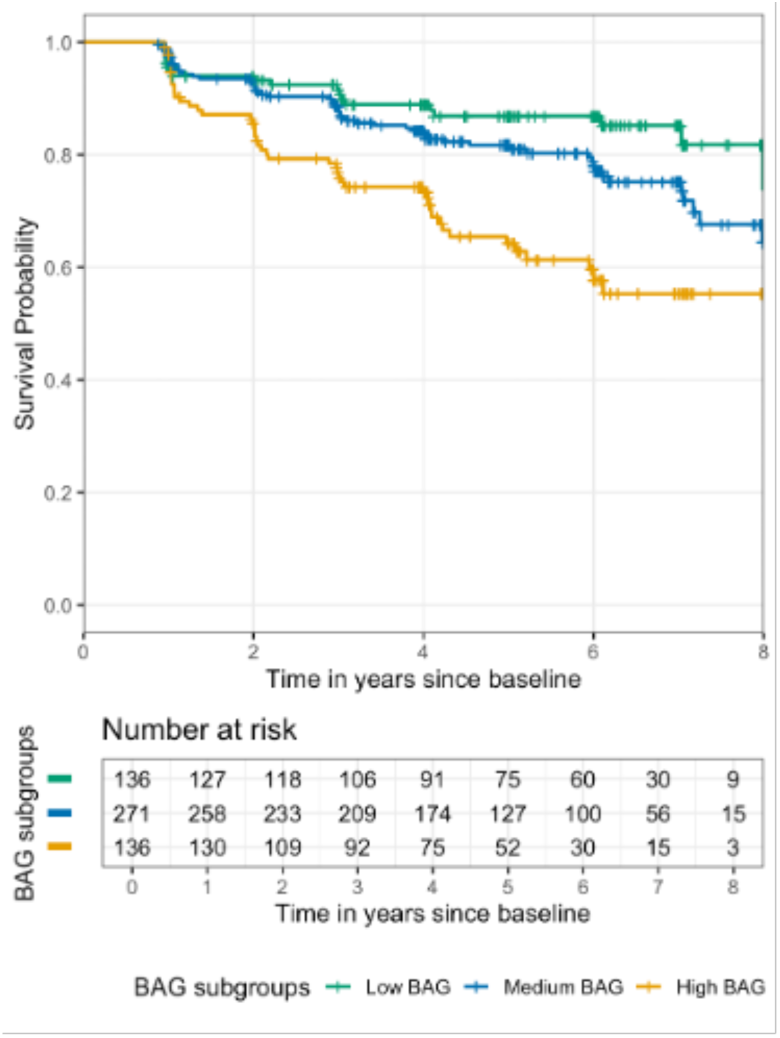
Kaplan-Meier survival curves for progression to MCI or MND according to BAG subgroups. Notes. Participants were stratified into low, medium, and high BAG groups based on quartiles of BAG distribution. Curves display the probability of remaining free of MCI/MND over 8 years from baseline, with tick marks indicating censoring events. The risk table shows the number of participants at risk at each timepoint. MND=Major Neurocognitive Disorder.

### BAG and lifestyle risk

Higher LIBRA scores were associated with higher BAG at baseline (+0·79 years per point; 95% CI 0·5 to 1·0; p<0·001), but not with longitudinal BAG change (time*LIBRA, p=0·51). The association was evident in the Aβ-group (+0·79 years per point; 95% CI 0·5 to 1·01; p<0·0001), attenuated in the Aβ+ group (+0·32; −0·05 to 0·69; p=0·10), and absent in the Aβ+T+ group (+0·01; −0·466 to 0·68; p=0·98), whereas a borderline association remained in the Aβ+T− group (+0·47; −0·00 to 0·94; p=0·05; **appendix p11**).

Mediation analysis showed that higher LIBRA scores were associated with poorer baseline cognition partly via higher BAG (indirect effect β=−0·02; 95% CI −0·03 to −0·01; 47% mediation) and steeper cognitive decline (β=−0·003; −0·005 to −0·002; 42% mediation) partly via higher BAG. Indirect effects were significant in the Aβ-subgroup but not in Aβ+, although direct effects of LIBRA on cognitive decline remained significant in the latter (**Figure 4**). Among CU subsample, LIBRA remained associated with BAG (β=0·64; 95% CI 0·37-0·88; specific to the Aβ-subgroup), and the indirect effect via BAG was not significant for baseline cognitive performance (β=−0·004; −0·008 to 0·000) but was significant for subsequent cognitive decline (β=−0·001; −0·002 to −0·000; appendix p12).

**Figure 4.**
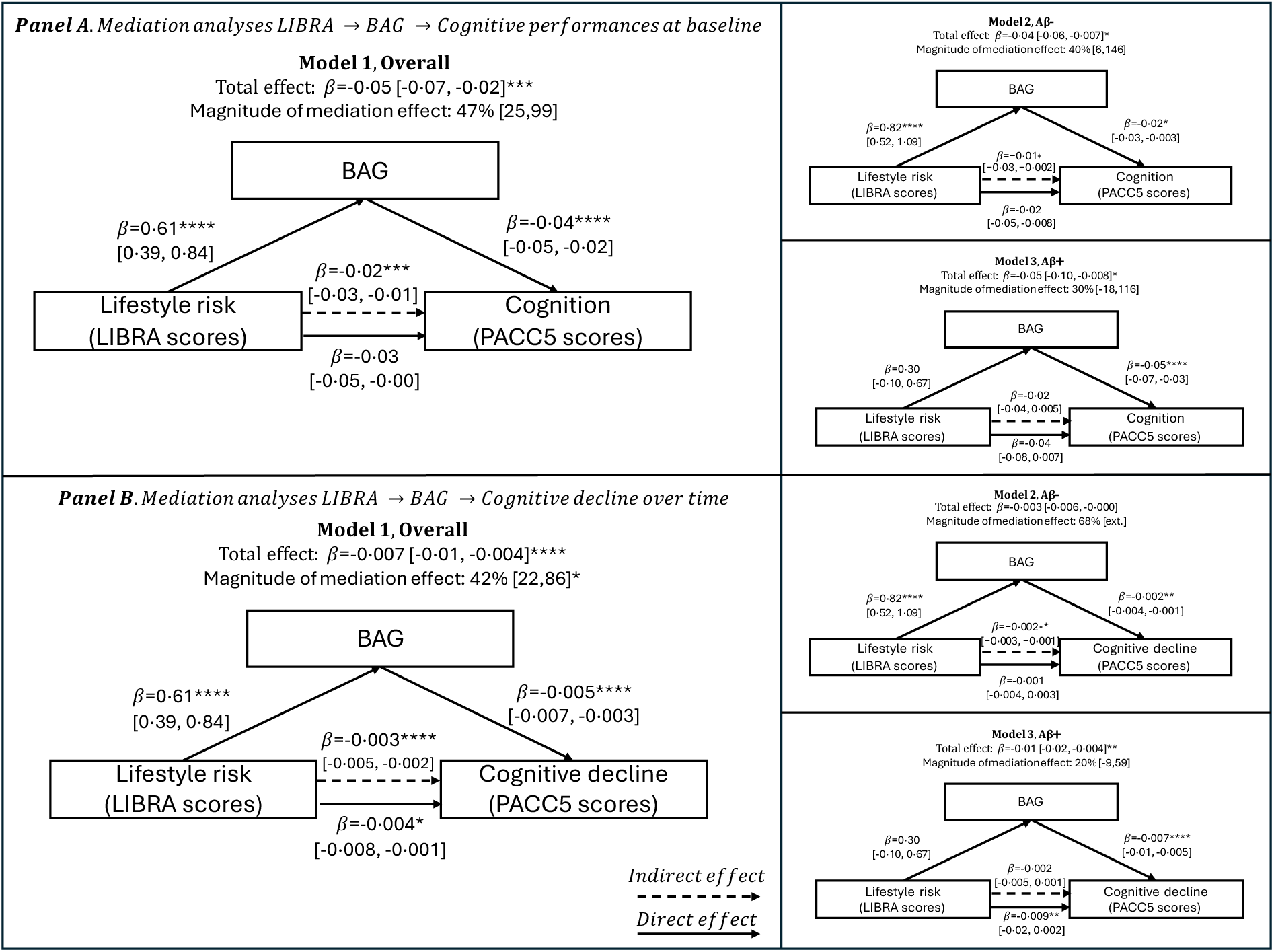
Mediation of the association between lifestyle risk (LIBRA) and cognition by brain age gap (BAG) Notes. Mediation models assessing whether BAG accounts for the association between lifestyle risk (LIBRA) and cognitive performance at baseline (panel A) or cognitive decline over time (panel B, based on PACC5 slopes). Models are shown for the overall sample (Model 1), amyloid-negative participants (Model 2, Aβ−), and amyloid-positive participants (Model 3, Aβ+). Solid arrows represent direct effects; dashed arrows represent indirect (mediated) effects. β coefficients are reported with 95% CIs. The magnitude of the mediation effect reflects the proportion of the total effect explained by BAG.

### Replication cohort

In ADNI, baseline BAG was also higher in Aβ+ and Aβ+T+ participants (Aβ+ vs Aβ-: +2·7 [0·6-4·7] years, p=0·01; Aβ+T+ vs Aβ-T−: +4·0 [1·0-6·9] years, p=0·004), compared to Aβ-[T−] participants. Longitudinally, BAG slopes differed by biomarker status, with accelerated increases in Aβ+ (+0·5 [0·1-0·8] years per year vs Aβ-, p=0·007) and most clearly in Aβ+T+ participants (+0·7 [0·2-1·2] years per year vs Aβ-T−, p=0·004; **appendix pp13-16**). Higher BAG was associated with lower cognitive performance and steeper cognitive decline, with stronger BAG-cognition slopes in biomarker-positive groups (**appendix pp17-18**). Baseline BAG also predicted clinical progression (HR 1·05, 95% CI 1·01-1·08, p=0·007), consistent with the main cohort analyses (**appendix p19)**.

## Discussion

This study shows that BAG captures stage-dependent changes in brain ageing across the AD continuum, reflecting a transition from modifiable risk-related brain ageing to pathology-driven structural decline.

Our findings confirm previous observations linking higher BAG to poorer cognition, increased risk of clinical progression, and sensitivity to AD-related neurodegeneration.^9,11,12^ These results support BAG as a global marker of brain ageing with clinical relevance.

Importantly, this study extends prior work by providing, to our knowledge, the first characterization of BAG across the full clinical and biological continuum of AD. BAG was already elevated in amyloid-positive patients with SCD, indicating that a measurable brain-age increase may emerge already in stage 2 of the AD continuum, preceding objective cognitive impairment.^25,26^ This finding may help explain inconsistent associations reported in CU participants,^13–15^ and suggests that Aβ+SCD represents a more informative stage than asymptomatic Aβ+ status alone for detecting early structural brain changes.

Longitudinally, BAG trajectories varied by baseline biomarker status, with the steepest increases observed in Aβ+T+ participants, consistent with evidence that tau pathology is a key driver of cortical atrophy and structural decline.^27^ Replication across DELCODE and ADNI supports the robustness of BAG across cohorts and biomarker modalities (fluid- and PET-based). Together, these findings support a continuum between healthy and pathological brain ageing,^9^ with progressive BAG divergence over time.

Higher BAG was associated with lower cognitive performance, steeper longitudinal cognitive decline, and higher risk of clinical progression independently of hippocampal volume, a key regional marker of AD-related neurodegeneration. This suggests that BAG captures a whole-brain signal not fully reflected by regional measures.^8,9,12^ Associations with progression remained after accounting for baseline cognition, indicating that BAG provides information beyond initial cognitive status. Rather than reflecting focal atrophy or baseline cognition alone, BAG appears to index a broader aspect of brain organisation relevant to clinical trajectories.

Associations were detectable even among CU participants in restricted analyses and were strongest in Aβ+T+ participants, further supporting sensitivity across disease stages and providing, to our knowledge, the first evidence that longitudinal BAG trajectories are already altered during the preclinical AD stage. In this context, BAG appears to complement, rather than replace, established regional structural markers such as hippocampal volume, which remain central outcomes in AD research.

The association between BAG and lifestyle-related dementia risk provides additional insight into the determinants of brain ageing. In DELCODE, higher LIBRA scores were associated with higher BAG primarily in biomarker-negative participants, with attenuation in Aβ+T− participants and no clear association in Aβ+T+ participants. This pattern suggests that modifiable risk factors contribute to early brain ageing before detectable AD pathology, whereas pathology-related mechanisms increasingly dominate once tau pathology is established. Mediation analyses indicated that BAG partly accounted for the association between lifestyle-related risk and cognition in the overall cohort, but not in biomarker-positive participants. These findings support a shift in the relative contribution of lifestyle-related and pathology-driven processes as disease progresses, suggesting that future prevention studies should consider biomarker status when evaluating lifestyle-related brain outcomes.^28^ Notably, hippocampal volume was not associated with lifestyle-related risk, further supporting that BAG captures global ageing-related effects beyond regional markers.

These findings have implications for research and clinical applications. As a scalable MRI-derived measure, BAG and their longitudinal trajectories complement molecular biomarkers by providing a global index of past and ongoing brain ageing which is sensitive to both, early risk-related burden and later disease processes. Within contemporary biomarker frameworks,^23^ BAG should not be viewed as a marker of AD pathology *per se*, but rather as an integrative measure of brain ageing at the intersection of modifiable risk exposure and neurodegeneration and may be conceptualized a “Core-1 adjacent” marker within the NIA-AA framework.^25^ Future studies should determine whether combining BAG with molecular biomarkers improves early risk stratification, prognostic accuracy, and longitudinal disease monitoring.

Several limitations should be noted. Tau classification partly relied on plasma p-tau_181_, which has lower specificity than tau-PET or newer plasma assays, although replication in ADNI supports robustness. Lifestyle-related risk was assessed cross-sectionally, limiting causal inference. Longitudinal BAG changes were modest, consistent with the slow pace of structural ageing. Finally, BAG estimates depend on modelling and acquisition factors, underscoring the need for methodological standardisation.^29^

In conclusion, BAG reflects the interplay between lifestyle-related burden and AD-related neurodegeneration across the disease continuum. By situating neurodegeneration within the broader biology of ageing, BAG may help identify when trajectories begin to diverge towards pathological decline.

## Contributors

EK contributed to the literature search, study conceptualization, data analysis, interpretation, and visualization, and writing and revision of the original draft. MW, GA, KP contributed to the study conceptualization, data analysis and interpretation, and revision of the original draft. SBE contributed to the study conceptualization and revision of the manuscript. All others co-authors contributed to the collection and preparation of DELCODE data and the review and revision of the manuscript. All authors had access to the data and had final responsibility for the decision to submit for publication.

## Data sharing

A request for access to the DELCODE data from qualified researchers can be submitted to the DELCODE steering committee (https://www.dzne.de/en/research/research-areas/clinical-research/for-researchers/).

## Acknowledgements

This work was funded by a Helmholtz Artificial Intelligence Cooperation Unit research grant awarded to Dr. Elizabeth Kuhn, Prof. Dr. Michael Wagner, Dr. Kaustbubh Patil and Prof. Dr. Simon B. Eickhoff.

