## Appendix for "MRI-derived brain aging trajectories across the Alzheimer’s continuum: links to cognition, pathology and lifestyle risk factors"

### Supplementary Online Content

*Notes· Left panels show baseline BAG distribution across clinical groups (A), clinical-biological AD subgroups (C), and A/T profiles (E)· Right panels (B, D, F) display corresponding individual longitudinal BAG trajectories; the dashed box indicates the baseline window (t=0)· Across groups, BAG increased stepwise with clinical severity, was higher in amyloid-positive symptomatic groups, and reached its highest values in Aβ+T1+ individuals· Abbreviations: CN=cognitively normal, SCD=subjective cognitive decline, MCI=mild cognitive impairment, DAT=Alzheimer's type dementia, BAG=brain-age gap, Aβ=β-amyloid, T=tau·* ........ 14

#### eMethods. Participants and Study Design, clinical follow-up data, and AD biomarkers.

##### 1. DELCODE (DZNE-Longitudinal Cognitive Impairment and Dementia Study).

###### 1.1. Participants:

The DELCODE study is a longitudinal, observational, memory clinic-based study conducted across nine sites in Germany, seven of which are linked to local university memory centers. Initiated in 2014 under the leadership of Prof. Dr. med. Frank Jessen, inclusion and exclusion criteria are described in Jessen et al., 2018.[1] The current study focuses on controls, first-degree relatives of Alzheimer's disease (AD) patients, and patients with subjective cognitive decline (SCD), mild cognitive impairment (MCI) or mild dementia of Alzheimer's type (DAT). Recruitment occurred through local newspaper advertisements and memory center referrals. All cognitively unimpaired (CU) participants scored better than 1·5 standard deviations below the age-, sex-, and education-adjusted norms on the CERAD neuropsychological battery (German norms available at www.memoryclinic.ch). SCD patients were additionally defined by subjectively reported cognitive decline, with concerns raised to memory center physicians, in line with criteria from previous studies.[2,3] Only participants with amnestic MCI and mild ADD (Mini-Mental State Examination [MMSE] ≥18, diagnosed according to the National Institute on Aging–Alzheimer's Association research criteria) were recruited in the MCI and DAT groups.[1,4,5] Eligible participants in the current study were aged 60-88, had at least 7 years of education, and no significant psychiatric, neurological, or substance abuse history. They also did not take psychoactive or anti-dementia medications. In total, 202 controls, 248 SCD patients, 93 MCI patients and 34 DAT were included.

###### 1.1.1. Clinical conversion to incident-MCI or dementia:

Consensus diagnoses of incident MCI were determined through a two-step review process, adapted from the Wisconsin Registry for Alzheimer's Prevention study and described in Stark et al, 2023.[6] In the first step, an algorithmic screening identified participants who were cognitively normal at baseline but showed potential cognitive decline at follow-up. The criteria for this included: (i) normative deficits (e.g., at least two cognitive tests ≥1·0 SD below the demographically adjusted mean, or one CERAD-NAB subtest ≥1·5 SD below the mean), (ii) individual cut-offs (e.g., CERAD-NAB word list delayed recall or FCSRT free recall ≥1·5 SD below the mean, FCSRT total recall ≤46 points, or MMSE ≤26 points), and (iii) reports from physicians or study partners (e.g., an increase in CDR global score, or FAQ score ≥4). In the second step, flagged cases were reviewed by a team of five neuropsychologists who assessed MCI conversion using established diagnostic criteria: (i) cognitive impairment (≥1·5 SD below the mean in at least two tests measuring the same ability), (ii) intra-individual cognitive decline (based on longitudinal test scores or participant/study partner reports), (iii) preserved functional abilities, and (iv) absence of major neurocognitive disorder (MND).[4,7] The review committee was blinded to baseline group assignments, biomarkers, imaging data, and genetic information.
Consensus diagnoses of incident dementia was diagnosed by the DELCODE study physicians based on established diagnostic criteria.[8-14] These diagnoses were additionally checked for inconsistencies by a review panel.

###### 1.2. AD biomarkers:

In the current study, amyloid and tau positivity were first assessed using cerebrospinal fluid (CSF) data, and plasma data when CSF was unavailable.
**CSF Biomarkers**: A subset of participants underwent lumbar puncture for CSF sampling (N=339, sampling rate: 58·8%). Amyloid beta (Aβ)42, Aβ40, and phospho-tau181 (p-tau181) levels were measured using standardized commercial kits: V-PLEX Aβ Peptide Panel 1 (6E10) Kit (K15200E, Mesoscale Diagnostics LLC, USA) and Innotest Phospho-Tau(181P) (81581; Fujirebio Germany GmbH). Assays were performed centrally, with independent reference samples used for quality control. Cut-offs were derived using Gaussian mixture modeling (R package flexmix, version 2.3-15)[15] and set at Aβ42/40 ≤ 0·08 and p-tau181 ≥ 73·65 pg/ml.[16]
**Plasma Aβ42/40:** Plasma samples (500 µL EDTA) were processed according to DELCODE standard operating procedures and stored at −80°C. AβX-40 and AβX-42 levels, along with the AβX-42/X-40 ratio, were measured using a semi-automated immunoprecipitation-immunoassay (IP-IA). The plasma Aβ immunoprecipitation was performed using a CyBio FeliX liquid-handling instrument (Roboscreen, Leipzig, Germany), followed by detection with the Mesoscale Discovery Aβ V-PLEX immunoassay (6E10). The cut-off for plasma Aβ42/40 was set at ≤ 0·106, as detailed in Vogelgsang et al., 2024.[17]
**Plasma pTau181**: Plasma levels of p-tau181 were measured using Simoa assays on Quanterix HD-1 and HD-X instruments (Quanterix, Billerica, MA) with pTau-181 Advantage version 2 kits, following the manufacturer's instructions. Assays were performed by the same operator, with two internal control samples assessed at the start and end of each run to ensure repeatability and monitor inter-assay variability.[18] The plasma p-tau181 cut-off for this study was determined via receiver operating characteristic (ROC) analysis, using CSF p-tau181 classification and the Youden Index to establish the threshold. The cut-off was set at ≥1·707 pg/mL, with an area under the curve (AUC) of 0·78 (95% CI: 0·73-0·83), based on 466 DELCODE participants, including MCI and demented patients (unpublished).

#### 2. ADNI (Alzheimer's Disease Neuroimaging Initiative).

In the current study, we utilized data from the ADNIMERGE dataset, extracted on September 21, 2023.

##### 2.1. Participants:

ADNI is an ongoing, longitudinal, multicenter study conducted in 63 sites across the USA and Canada. The ADNI was launched in 2003 as a public-private partnership, led by Principal Investigator Michael W. Weiner, MD. The primary goal of ADNI has been to test whether serial magnetic resonance imaging (MRI), positron emission tomography (PET), other biological markers, and clinical and neuropsychological assessment can be combined to measure the progression of mild cognitive impairment (MCI) and early Alzheimer's disease (AD). Further details can be found in Weiner et al,[19] and inclusion and exclusion criteria have been described at http://adni.loni.usc.edu/. Briefly, participants included in the current study were all aged between 55-88 years, had at least 7 years of education, and had no clinically significant psychiatric (including alcohol or drug abuse) or neurologic disease. All cognitively unimpaired (CU) participants had MMSE[20] scores of 24 or greater (total possible range: 0-30) and Clinical Dementia Rating (CDR) of 0.0 (total possible range 0-3, with higher scores indicating worse functioning)[21], are not depressed, did not have mild cognitive impairment or MND, have preserved activities of daily living (FAQ≤9), and had scores on delayed recall of one paragraph from Wechsler Memory Scale Logical Memory II within the norm according to their level of education (≥9 for 16 or more years of education, ≥5 for 8-15 years of education, ≥3 for 0-7 years of education). Part of those participants (N=145 [51·8%]) had a significant subjective cognitive decline, defined by the presence of a significant subjective memory concern reported by subject, informant, or clinician, CCI score ≥16 (based on first 12 questions).[22] Patients with MCI had CDR of 0.5 and fulfilled the standard diagnostic criteria for MCI (preserved activities of daily living)[23]; while patients with AD dementia (DAT patients) had CDR scores of 1.0 or greater, and fulfilled the standard diagnostic criteria of NINCDS-ADRDA for probable AD (dementia, progressive impairment, and absence of other diseases capable of producing dementia)[8].

**Clinical Conversion to Incident MCI or dementia**: To determine the presence of a clinical conversion during the follow-up period, we used an algorithm based on the "DX" (diagnosis) column from the ADNIMERGE dataset. Individuals diagnosed with mild cognitive impairment (MCI) or dementia at any point during follow-up were considered converters to incident MCI or dementia from the time of diagnosis. However, individuals with reversible MCI, defined as those who had an inconsistent MCI diagnosis, followed by a return to normal cognition in subsequent assessments (with at least three total assessments), were considered stable and not counted as converters.

##### 2.2. AD biomarkers:

**Aβ-PET biomarkers:** The ADNI FreeSurfer 5.3 pipeline yields a global standard uptake value ratio (SUVR) measure representing a non-weighted average of radiotracer retention in four FreeSurfer-defined regions (frontal, anterior/posterior cingulate, lateral parietal, and lateral temporal cortices) normalized to whole cerebellum. To directly convert [18F]-Florbetapir- and [18F]-Florbetaben-PET data to centiloid values, equations describe in ADNI guidelines were used (see details here).
Centiloids were calculated as follows:

- [18F]-Florbetapir-PET: CL=(196·9×SUVR)-196·03. The amyloid positivity threshold was set at 22·529, corresponding to 1·11 SUVR (see also UCBERKELEY_AV45_Methods_11.15.2021.pdf).[24,25]
- [18F]-Florbetaben-PET: CL=(159·08×SUVR)-151·65. The amyloid positivity threshold was set at 20·1564, corresponding to 1·08 SUVR (see also UCBerkeley_FBB_Methods_11.15.2021.pdf).[26]

**Tau-PET biomarker:** The processing of [18F]-Flortaucipir PET images is summarized as follows (please see here for details). Each PET scan is co-registered to the closest available bias-corrected T1-weighted MRI in native space, using FreeSurfer (v7.1.1) for anatomical accuracy. This registration allows the calculation of the mean flortaucipir uptake within predefined regions of interest (ROIs). For the current study, tau positivity is defined based on the standardized uptake value ratio (SUVR) in the entorhinal cortex (corresponding to Braak stage 1). A positivity threshold of SUVR≥1·2 was used, as established in a previously published paper.[27]

**eTable 1.** Model performances of BrainageR in healthy controls participants from the DELCODE study.

| Model | MAE | RMSE | R2 | r |
|---|---|---|---|---|
| **Direct prediction performances** | | | | |
| brainageR (raw) | 14·55 | 16·78 | 0·31 | 0·56 |
| brainageR (age-corrected) | 7·54 | 9·22 | 0·25 | 0·50 |
| **Linear calibration model** (Chronological Age ~ MRI-derived predicted age) | | | | |
| brainageR (raw) | 3·41 | 4·30 | 0·36 | 0·60 |
| brainageR (age-corrected) | 3·56 | 4·49 | 0·30 | 0·55 |

#### eTable 2. Longitudinal patterns of Brain Age Gap (BAG) across clinical groups.

| | Overall | | |
|---|---|---|---|
| **Model 1 - Fixed Effect[a]** | Est· | SE | P |
| (Intercept) | 9·70 | 4·74 | **0·041** |
| Time | 1·71 | 0·75 | **0·024** |
| Group | | | **<0·001** |
| SCD | 1·45 | 0·77 | |
| MCI | 4·28 | 1·04 | |
| DAT | 9·48 | 1·55 | |
| Time:Group | | | 0·431 |
| SCD | 0·13 | 0·11 | |
| MCI | -0·11 | 0·17 | |
| DAT | 0·08 | 0·33 | |
| **Slopes** | Est· | 95% CI | $P_{adj}$ |
| CN | -0·03 | -0·30, 0·25 | 0·820 |
| SCD | 0·11 | -0·16, 0·37 | 0·653 |
| MCI | -0·13 | -0·48, 0·21 | 0·653 |
| DAT | 0·05 | -0·55, 0·66 | 0·820 |
| **Pairwise comparisons (BL)** | Est· | 95% CI | $P_{adj}$ |
| CN / SCD | -1·67 | -3·71, 0·38 | **0·031** |
| CN / MCI | -4·10 | -6·87, -1·33 | **<0·001** |
| CN / DAT | -9·61 | -13·81, -5·42 | **<0·001** |
| SCD / MCI | -2·43 | -5·06, 0·19 | **0·017** |
| SCD / DAT | -7·94 | -12·05, -3·84 | **<0·001** |
| MCI / DAT | -5·51 | -9·91, -1·11 | **0·001** |
| **Pairwise comparisons (Slopes)** | Est· | 95% CI | $P_{adj}$ |
| CN / SCD | -0·13 | -0·43, 0·17 | 0·732 |
| CN / MCI | 0·11 | -0·34, 0·56 | 0·874 |
| CN / DAT | -0·08 | -0·95, 0·78 | 0·874 |
| SCD / MCI | 0·24 | -0·19, 0·67 | 0·732 |
| SCD / DAT | 0·05 | -0·80, 0·91 | 0·874 |
| MCI / DAT | -0·19 | -1·10, 0·72 | 0·874 |

*Abbreviations: CN=cognitively normal, SCD=subjective cognitive decline, MCI=mild cognitive impairment, DAT=Alzheimer's type major neurocognitive disorder, BAG=brain-age gap, Aβ=β-amyloid, T=tau.*

**eTable 3. Longitudinal patterns of Brain Age Gap (BAG) across clinical-biological AD construct (CN Aβ- vs others Aβ+ clinical groups).**

| | Overall | | |
|---|---|---|---|
| **Model 1 - Fixed Effect[a]** | Est· | SE | P |
| (Intercept) | 10·59 | 6·12 | *0·085* |
| Time | 2·63 | 0·92 | **0·005** |
| Group | | | **<0·001** |
| CN·Aβ+ | -1·25 | 1·35 | |
| SCD·Aβ+ | 2·16 | 1·14 | |
| MCI·Aβ+ | 3·15 | 1·36 | |
| DAT·Aβ+ | 9·57 | 1·76 | |
| Time:Group | | | *0·058* |
| CN·Aβ+ | -0·25 | 0·19 | |
| SCD·Aβ+ | 0·36 | 0·16 | |
| MCI·Aβ+ | 0·10 | 0·21 | |
| DAT·Aβ+ | 0·22 | 0·34 | |
| **Slopes** | Est· | 95% CI | $P_{adj}$ |
| CN·Aβ+ | -0·33 | -0·73, 0·07 | 0·102 |
| SCD·Aβ+ | 0·27 | -0·07, 0·61 | 0·102 |
| MCI·Aβ+ | 0·02 | -0·41, 0·44 | 0·917 |
| DAT·Aβ+ | 0·13 | -0·56, 0·82 | 0·775 |
| **Pairwise comparisons (BL)** | Est· | 95% CI | $P_{adj}$ |
| CN·Aβ- / CN·Aβ+ | 1·66 | -2·16, 5·49 | 0·245 |
| CN·Aβ- / SCD·Aβ+ | -2·75 | -5·97, 0·46 | **0·020** |
| CN·Aβ- / MCI·Aβ+ | -3·33 | -7·17, 0·52 | **0·020** |
| CN·Aβ- / DAT·Aβ+ | -9·94 | -14·97, -4·90 | **<0·001** |
| CN·Aβ+ / SCD·Aβ+ | -4·41 | -8·51, -0·32 | **0·004** |
| CN·Aβ+ / MCI·Aβ+ | -4·99 | -9·63, -0·35 | **0·004** |
| CN·Aβ+ / DAT·Aβ+ | -11·60 | -17·26, -5·94 | **<0·001** |
| SCD·Aβ+ / MCI·Aβ+ | -0·57 | -4·51, 3·36 | 0·681 |
| SCD·Aβ+ / DAT·Aβ+ | -7·18 | -12·25, -2·11 | **<0·001** |
| MCI·Aβ+ / DAT·Aβ+ | -6·61 | -11·99, -1·24 | **0·001** |
| **Pairwise comparisons (Slopes)** | Est· | 95% CI | $P_{adj}$ |
| CN·Aβ- / CN·Aβ+ | 0·25 | -0·28, 0·77 | 0·412 |
| CN·Aβ- / SCD·Aβ+ | -0·36 | -0·82, 0·11 | 0·156 |
| CN·Aβ- / MCI·Aβ+ | -0·10 | -0·69, 0·49 | 0·754 |
| CN·Aβ- / DAT·Aβ+ | -0·22 | -1·19, 0·75 | 0·749 |
| CN·Aβ+ / SCD·Aβ+ | -0·60 | -1·18, -0·02 | **0·038** |
| CN·Aβ+ / MCI·Aβ+ | -0·35 | -1·04, 0·34 | 0·412 |
| CN·Aβ+ / DAT·Aβ+ | -0·46 | -1·50, 0·57 | 0·412 |
| SCD·Aβ+ / MCI·Aβ+ | 0·25 | -0·37, 0·87 | 0·418 |
| SCD·Aβ+ / DAT·Aβ+ | 0·14 | -0·85, 1·12 | 0·754 |
| MCI·Aβ+ / DAT·Aβ+ | -0·12 | -1·15, 0·92 | 0·754 |

**eTable 4.** Longitudinal patterns of Brain Age Gap (BAG) across amyloid status (model 1) or A/T status (model 2, excluding Aβ-.T+).

| | **Overall** | | | **CU** | | |
|---|---|---|---|---|---|---|
| **Model 1 - Fixed Effect[a]** | Est· | SE | P | Est· | SE | P |
| (Intercept) | 7·29 | 4·88 | 0·136 | 3·97 | 5·55 | 0·475 |
| Time | 1·75 | 0·75 | **0·020** | 1·06 | 0·80 | 0·186 |
| Group [Aβ+] | 1·34 | 0·75 | **0·043** | 0·31 | 0·85 | 0·718 |
| Time:Group [Aβ+] | 0·10 | 0·11 | 0·355 | 0·04 | 0·12 | 0·729 |
| **Model 2 - Fixed Effect[a]** | Est· | SE | P | Est· | SE | P |
| (Intercept) | 7·26 | 5·09 | 0·154 | 3·96 | 5·68 | 0·487 |
| Time | 2·12 | 0·77 | **0·006** | 1·32 | 0·83 | 0·114 |
| Group | | | 0·203 | | | 0·251 |
| Aβ+·T- | 1·13 | 0·86 | | 0·78 | 0·94 | |
| Aβ+·T+ | 1·68 | 1·07 | | -1·73 | 1·43 | |
| Time:Group | | | **0·003** | | | *0·078* |
| Aβ+·T- | -0·07 | 0·13 | | -0·05 | 0·13 | |
| Aβ+·T+ | 0·55 | 0·18 | | 0·49 | 0·23 | |
| **Slopes** | Est· | 95% CI | $P_{adj}$ | Est· | 95% CI | $P_{adj}$ |
| Aβ-·T- | -0·16 | -0·35, 0·03 | **0·047** | -0·15 | -0·38, 0·09 | 0·130 |
| Aβ+·T- | -0·23 | -0·45, -0·01 | **0·017** | -0·20 | -0·45, 0·06 | *0·097* |
| Aβ+·T+ | 0·39 | 0·11, 0·67 | **0·002** | 0·34 | -0·02, 0·70 | *0·073* |
| **Pairwise comparisons (BL)** | Est· | 95% CI | Padj | Est· | 95% CI | $P_{adj}$ |
| Aβ-·T- / Aβ+·T- | -1·02 | -3·08, 1·05 | 0·236 | -0·69 | -2·95, 1·56 | 0·540 |
| Aβ-·T- / Aβ+·T+ | -2·61 | -5·19, -0·02 | **0·047** | 0·88 | -2·57, 4·32 | 0·540 |
| Aβ+·T- / Aβ+·T+ | -1·59 | -4·35, 1·18 | 0·236 | 1·57 | -2·08, 5·22 | 0·540 |
| **Pairwise comparisons (Slopes)** | Est· | 95% CI | Padj | Est· | 95% CI | $P_{adj}$ |
| Aβ-·T- / Aβ+·T- | 0·07 | -0·24, 0·37 | 0·591 | 0·05 | -0·27, 0·37 | 0·713 |
| Aβ-·T- / Aβ+·T+ | -0·55 | -0·98, -0·13 | **0·003** | -0·49 | -1·04, 0·07 | *0·052* |
| Aβ+·T- / Aβ+·T+ | -0·62 | -1·08, -0·16 | **0·003** | -0·54 | -1·12, 0·04 | *0·052* |

**eTable 5. Longitudinal cognitive performances (PACC5 scores) according to BAG at baseline.**

| | **Adj· demographics** | | | **Adj for hippocampal volume** | | |
|---|---|---|---|---|---|---|
| **Model 1· Overall - Fixed Effect** | Est· | SE | P | Est· | SE | P |
| (Intercept) | 2·76 | 0·62 | **<0·001** | -1·11 | 0·87 | 0·199 |
| Time | 0·40 | 0·13 | **0·002** | -0·16 | 0·17 | 0·352 |
| BAG | -0·04 | 0·005 | **<0·001** | -0·02 | 0·006 | **<0·001** |
| Time:BAG | -0·006 | 0·001 | **<0·001** | -0·003 | 0·001 | **0·005** |
| **Model 2· Overall - Fixed Effect** | Est· | SE | P | Est· | SE | P |
| (Intercept) | 2·19 | 0·60 | **<0·001** | -1·09 | 0·84 | 0·196 |
| Time | 0·30 | 0·12 | **0·014** | -0·15 | 0·17 | 0·368 |
| BAG | -0·03 | 0·006 | **<0·001** | -0·01 | 0·007 | *0·090* |
| BAG:Group [Aβ+] | -0·02 | 0·009 | **0·030** | -0·02 | 0·009 | **0·012** |
| Time:BAG | -0·003 | 0·001 | **0·010** | -0·001 | 0·001 | 0·384 |
| Time:BAG:Group [Aβ+] | -0·005 | 0·002 | **0·005** | -0·005 | 0·002 | **0·003** |
| **Model 3· Overall - Fixed Effect** | Est· | SE | P | Est· | SE | P |
| (Intercept) | 1·86 | 0·59 | **0·002** | -1·14 | 0·81 | 0·160 |
| Time | 0·17 | 0·12 | 0·157 | -0·22 | 0·16 | 0·169 |
| BAG | -0·02 | 0·006 | **<0·001** | -0·009 | 0·007 | 0·175 |
| BAG:Group | | | **0·005** | | | **0·002** |
| Aβ+T- | -0·01 | 0·010 | | -0·01 | 0·010 | |
| Aβ+T+ | -0·04 | 0·013 | | -0·04 | 0·012 | |
| Time:BAG | -0·003 | 0·001 | **0·022** | -0·001 | 0·001 | 0·419 |
| Time:BAG:Group | | | **0·005** | | | **0·004** |
| Aβ+T- | -0·004 | 0·002 | | -0·004 | 0·002 | |
| Aβ+T+ | -0·009 | 0·003 | | -0·009 | 0·003 | |
| **Pairwise comparisons (BL)** | Est· | 95% CI | $P_{adj}$ | Est· | 95% CI | $P_{adj}$ |
| Aβ-·T- / Aβ+·T- | 0·02 | -0·01, 0·05 | 0·10 | 0·02 | -0·008, 0·05 | *0·076* |
| Aβ-·T- / Aβ+·T+ | 0·06 | 0·02, 0·10 | **<0·001** | 0·07 | 0·03, 0·10 | **<0·001** |
| Aβ+·T- / Aβ+·T+ | 0·04 | -0·001, 0·08 | **0·03** | 0·04 | -0·003, 0·09 | **0·016** |
| **Pairwise comparisons (Slopes)** | Est· | 95% CI | $P_{adj}$ | Est· | 95% CI | $P_{adj}$ |
| Low (-7·6 y) | | | | | | |
| Aβ−T− / Aβ+T+ | 0·19 | 0·04, 0·34 | **<0·001** | 0·17 | 0·02, 0·32 | **<0·001** |
| Aβ−T− / Aβ+T- | 0·05 | −0·04, 0·14 | *0·092* | 0·04 | −0·05, 0·13 | 0·146 |
| Aβ+T− / Aβ+T+ | 0·14 | −0·02, 0·30 | **0·008** | 0·13 | −0·03, 0·29 | **0·013** |
| Mean (+2·0 y) | | | | | | |
| Aβ−T− / Aβ+T+ | 0·28 | 0·18, 0·38 | **<0·001** | 0·26 | 0·16, 0·36 | **<0·001** |
| Aβ−T− / Aβ+T- | 0·09 | 0·02, 0·15 | **<0·001** | 0·08 | 0·02, 0·14 | **<0·001** |
| Aβ+T− / Aβ+T+ | 0·19 | 0·08, 0·30 | **<0·001** | 0·18 | 0·08, 0·29 | **<0·001** |
| High (+11·5 y) | | | | | | |
| Aβ−T− / Aβ+T+ | 0·36 | 0·24, 0·49 | **<0·001** | 0·35 | 0·23, 0·48 | **<0·001** |
| Aβ−T− / Aβ+T- | 0·12 | 0·04, 0·20 | **<0·001** | 0·12 | 0·04, 0·20 | **<0·001** |
| Aβ+T− / Aβ+T+ | 0·24 | 0·11, 0·37 | **<0·001** | 0·23 | 0·10, 0·37 | **<0·001** |
| Low vs High | | | | | | |
| Aβ−T− | **0.05** | −0.02, 0.12 | **0.025** | **0.02** | −0.06, 0.09 | 0.431 |
| Aβ+T− | **0.12** | 0.02, 0.23 | **<0.001** | **0.09** | −0.02, 0.20 | **0.009** |
| Aβ+T+ | **0.23** | 0.04, 0.41 | **<0.001** | **0.20** | 0.01, 0.38 | **0.001** |

**eTable 6. Longitudinal cognitive performances (PACC5 scores) according to BAG at baseline in CU subsample.**

| | **Adj· demographics** | | | **Adj for hippocampal volume** | | |
|---|---|---|---|---|---|---|
| **Model 1· Overall - Fixed Effect** | Est· | SE | P | Est· | SE | P |
| (Intercept) | 1·79 | 0·37 | **<0·001** | -1·06 | 0·52 | 0·042 |
| Time | 0·14 | 0·09 | 0·139 | 0·16 | 0·13 | 0·208 |
| BAG | -0·007 | 0·003 | **0·033** | -0·004 | 0·003 | 0·259 |
| Time:BAG | -0·002 | 0·001 | **0·003** | -0·002 | 0·0008 | **0·005** |
| **Model 2· Overall - Fixed Effect** | Est· | SE | P | Est· | SE | P |
| (Intercept) | 1·71 | 0·37 | **<0·001** | 1·05 | 0·52 | **0·04** |
| Time | 0·12 | 0·09 | 0·199 | 0·16 | 0·12 | 0·199 |
| BAG | -0·003 | 0·004 | 0·399 | -0·001 | 0·004 | 0·891 |
| BAG:Group [Aβ+] | -0·010 | 0·006 | *0·073* | -0·010 | 0·006 | *0·072* |
| Time:BAG | -0·001 | 0·001 | 0·111 | -0·01 | 0·001 | *0·099* |
| Time:BAG:Group [Aβ+] | -0·003 | 0·001 | *0·059* | -0·003 | 0·001 | *0·054* |
| **Model 3· Overall - Fixed Effect** | Est· | SE | P | Est· | SE | P |
| (Intercept) | 1·71 | 0·38 | **<0·001** | 0·90 | 0·53 | *0·09* |
| Time | 0·04 | 0·09 | 0·655 | 0·08 | 0·12 | 0·51 |
| BAG | -0·003 | 0·004 | 0·373 | -0·0003 | 0·004 | 0·94 |
| BAG:Group | | | 0·100 | | | *0·09* |
| Aβ+T- | -0·007 | 0·007 | | -0·006 | 0·006 | |
| Aβ+T+ | -0·020 | 0·010 | | -0·02 | 0·01 | |
| Time:BAG | -0·002 | 0·001 | *0·065* | -0·002 | 0·001 | *0·059* |
| Time:BAG:Group | | | **0·027** | | | **0·026** |
| Aβ+T- | -0·002 | 0·001 | | -0·002 | 0·001 | |
| Aβ+T+ | -0·007 | 0·003 | | -0·007 | 0·003 | |
| **Pairwise comparisons (BL)** | Est· | 95% CI | $P_{adj}$ | Est· | 95% CI | $P_{adj}$ |
| Aβ-·T- / Aβ+·T- | 0·01 | -0·006, 0·03 | 0·11 | 0·01 | -0·006, 0·03 | *0·076* |
| Aβ-·T- / Aβ+·T+ | 0·04 | 0·01, 0·06 | **0·002** | 0·04 | 0·01, 0·06 | **0·002** |
| Aβ+·T- / Aβ+·T+ | 0·03 | -0·003, 0·05 | **0·048** | 0·03 | -0·002, 0·05 | **0·041** |
| **Pairwise comparisons (Slopes)** | Est· | 95% CI | $P_{adj}$ | Est· | 95% CI | $P_{adj}$ |
| Low (-8·3/-8·1 y) | | | | | | |
| Aβ−T− / Aβ+T+ | 0·11 | −0·02, 0·24 | **0·011** | 0·11 | -0·01, 0·23 | **0·005** |
| Aβ−T− / Aβ+T- | 0·07 | −0·09, 0·23 | 0·20 | 0·03 | -0·04, 0·10 | 0·146 |
| Aβ+T− / Aβ+T+ | 0·08 | −0·05, 0·21 | *0·091* | 0·08 | -0·05, 0·21 | *0·069* |
| Mean (+1·0/+1·2 y) | | | | | | |
| Aβ−T− / Aβ+T+ | 0·19 | 0·09, 0·28 | **<0·001** | 0·18 | 0·09, 0·26 | **<0·001** |
| Aβ−T− / Aβ+T- | 0·15 | 0·01, 0·29 | **<0·001** | 0·05 | 0·01, 0·10 | **0·001** |
| Aβ+T− / Aβ+T+ | 0·12 | 0·03, 0·22 | **<0·001** | 0·12 | 0·04, 0·21 | **<0·001** |
| High (+10·3/+10·5 y) | | | | | | |
| Aβ−T− / Aβ+T+ | 0·25 | 0·14, 0·37 | **<0·001** | 0·24 | 0·13, 0·35 | **<0·001** |
| Aβ−T− / Aβ+T- | 0·24 | 0·12, 0·35 | **<0·001** | 0·07 | 0·01, 0·13 | **<0·001** |
| Aβ+T− / Aβ+T+ | 0·17 | 0·05, 0·29 | **<0·001** | 0·17 | 0·05, 0·29 | **<0·001** |
| Low vs High | | | | | | |
| Aβ−T− | **0.028** | −0.02, 0.08 | *0.076* | **0.030** | −0.02, 0.08 | *0.069* |
| Aβ+T− | **0.064** | −0.02, 0.14 | **0.017** | **0.067** | −0.02, 0.15 | **0.015** |
| Aβ+T+ | **0.156** | −0.001, 0.31 | **0.003** | **0.158** | 0.001, 0.32 | **0.003** |

#### eTable 7· Risk of clinical progression to incident mild cognitive impairment (iMCI) or major neurocognitive disorder according to BAG·

| | iMCI_Dem | | | iMCI | | | iDem | | |
|---|---|---|---|---|---|---|---|---|---|
| **Model 1** | $N_{obs[event]}$ | HR [95% CI] | P | $N_{obs[event]}$ | HR [95% CI] | P | $N_{obs[event]}$ | HR [95% CI] | P |
| BAG | 543 [123] | 1·05 [1·03, 1·08] | **<0·001** | 449 [88] | 1·05 [1·02, 1·08] | **<0·001** | 543 [42] | 1·09 [1·04, 1·13] | **<0·001** |
| **Model 1 * Aβ** | $N_{obs[event]}$ | HR [95% CI] | P | $N_{obs[event]}$ | HR [95% CI] | P | $N_{obs[event]}$ | HR [95% CI] | P |
| BAG | 543 [123] | 1·05 [1·02, 1·09] | **0·002** | 449 [88] | 1·05 [1·01, 1·09] | **0·007** | 543 [42] | 1·17 [1·04, 1·13] | **0·004** |
| BAG * Amyloid status [Aβ+] | 543 [69] | 1·00 [0·96, 1·04] | 0·90 | 449 [40] | 1·00 [0·95, 1·05] | 0·90 | 543 [36] | 0·91 [0·82, 1·01] | *0·09* |
| **Model 1 adj· Aβ** | $N_{obs[event]}$ | HR [95% CI] | P | $N_{obs[event]}$ | HR [95% CI] | P | $N_{obs[event]}$ | HR [95% CI] | P |
| BAG | 543 [123] | 1·05 [1·03, 1·07] | **<0·001** | 449 [88] | 1·05 [1·02, 1·08] | **<0·001** | 543 [42] | 1·08 [1·03, 1·12] | **<0·001** |
| Baseline amyloid status [Aβ+] | 543 [69] | 2·30 [1·58, 3·36] | **<0·001** | 449 [40] | 1·77 [1·13, 2·77] | **0·013** | 543 [36] | 10·30 [4·23, 25·10] | **<0·001** |
| **Model 1 * AβT** | $N_{obs[event]}$ | HR [95% CI] | P | $N_{obs[event]}$ | HR [95% CI] | P | $N_{obs[event]}$ | HR [95% CI] | P |
| BAG | 515 [118] | 1·06 [1·02, 1·09] | **0·001** | 412 [84] | 1·05 [1·02, 1·09] | **0·005** | 515 [41] | 1·19 [1·06, 1·33] | **0·003** |
| BAG * Ab+T- status | 515 [35] | 1·00 [0·95, 1·05] | 0·90 | 412 [22] | 1·00 [0·94, 1·06] | 0·90 | 515 [16] | 0·91 [0·80, 1·03] | 0·14 |
| BAG * Ab+T+ status | 515 [34] | 0·99 [0·95, 1·04] | 0·80 | 412 [18] | 1·01 [0·95, 1·07] | 0·80 | 515 [20] | 0·90 [0·80, 1·02] | *0·08* |
| **Model 1 adj· AβT** | $N_{obs[event]}$ | HR [95% CI] | P | $N_{obs[event]}$ | HR [95% CI] | P | $N_{obs[event]}$ | HR [95% CI] | P |
| BAG | 515 [118] | 1·05 [1·03, 1·08] | **<0·001** | 412 [84] | 1·05 [1·02, 1·09] | **0·005** | 515 [41] | 1·09 [1·04, 1·13] | **<0·001** |
| BAG * Ab+T- status | 515 [35] | 1·65 [1·05, 2·58] | 0·03 | 412 [22] | 1·00 [0·94, 1·06] | 0·90 | 515 [16] | 7·11 [2·56, 19·08] | **<0·001** |
| BAG * Ab+T+ status | 515 [34] | 4·87 [3·00, 7·88] | **<0·001** | 412 [18] | 1·01 [0·95, 1·07] | 0·80 | 515 [20] | 31·90 [11·10, 91·70] | **<0·001** |
| **Model 1 adj· hippocampal volume** | $N_{obs[event]}$ | HR [95% CI] | P | $N_{obs[event]}$ | HR [95% CI] | P | $N_{obs[event]}$ | HR [95% CI] | P |
| BAG | 543 [123] | 1·02 [1·00, 1·05] | *0·056* | 543 [88] | 1·03 [1·00, 1·06] | *0·054* | 543 [42] | 1·04 [0·99, 1·08] | 0·13 |
| Baseline hippocampal volume | 543 [123] | 0·52 [0·40, 0·68] | **<0·001** | 543 [88] | 0·57 [0·41, 0·80] | **<0·001** | 543 [42] | 0·28 [0·17, 0·45] | **<0·001** |
| **Model 1 adj· Pacc5** | $N_{obs[event]}$ | HR [95% CI] | P | $N_{obs[event]}$ | HR [95% CI] | P | $N_{obs[event]}$ | HR [95% CI] | P |
| BAG | 543 [123] | 1·03 [1·00, 1·05] | **0·018** | 543 [88] | 1·04 [1·01, 1·06] | **0·007** | 543 [42] | 1·04 [1·00, 1·09] | **0·054** |
| Baseline Pacc5 | 543 [123] | 0·41 [0·34, 0·50] | **<0·001** | 543 [88] | 0·22 [0·16, 0·30] | **<0·001** | 543 [42] | 0·22 [0·16, 0·30] | **<0·001** |

#### eTable 8· Longitudinal patterns of Brain Age Gap (BAG) across LIBRA·

| | Overall | | | CU | | |
|---|---|---|---|---|---|---|
| **Model 1 - Fixed Effect[a]** | Est. | SE | P | Est· | SE | P |
| (Intercept) | -15·57 | 5·13 | **0·002** | -23·31 | 5·81 | **<0·001** |
| Time | 1·49 | 0·71 | **0·035** | 0·79 | 0·75 | 0·30 |
| LIBRA | 0·79 | 0·13 | **<0·001** | 0·87 | 0·15 | **<0·001** |
| Time:LIBRA | 0·012 | 0·018 | 0·51 | 0·015 | 0·019 | 0·44 |
| **Aβ- participants** | Est. | SE | P | Est· | SE | P |
| (Intercept) | 3·03 | 5·93 | 0·61 | 0·38 | 6·55 | 0·95 |
| Time | 0·35 | 0·88 | 0·69 | -0·05 | 0·92 | 0·96 |
| LIBRA | 0·79 | 0·14 | **<0·001** | 0·83 | 0·16 | **<0·001** |
| Time:LIBRA | -0·003 | 0·022 | 0·88 | -0·007 | 0·02 | 0·76 |
| **Aβ+ participants** | Est. | SE | P | Est· | SE | P |
| (Intercept) | 8·26 | 8·36 | 0·32 | 3·22 | 10·33 | 0·76 |
| Time | 4·35 | 1·40 | **0·002** | 3·13 | 1·61 | *0·05* |
| LIBRA | 0·32 | 0·19 | 0·10 | 0·21 | 0·23 | 0·37 |
| Time:LIBRA | 0·032 | 0·032 | 0·31 | 0·05 | 0·04 | 0·17 |
| **Aβ-T- participants** | Est. | SE | P | Est· | SE | P |
| (Intercept) | 1·62 | 6·29 | 0·80 | 1·51 | 6·79 | 0·82 |
| Time | 0·42 | 0·93 | 0·65 | -0·04 | 0·98 | 0·97 |
| LIBRA | 0·74 | 0·15 | **<0·001** | 0·77 | 0·16 | **<0·001** |
| Time:LIBRA | -0·012 | 0·023 | 0·59 | -0·02 | 0·02 | 0·154 |
| **Aβ+T- participants** | Est. | SE | P | Est· | SE | P |
| (Intercept) | -1·69 | 10·84 | 0·88 | -3·60 | 12·38 | 0·77 |
| Time | 3·76 | 1·48 | **0·011** | 2·92 | 1·71 | *0·09* |
| LIBRA | 0·47 | 0·24 | **0·049** | 0·22 | 0·27 | 0·43 |
| Time:LIBRA | 0·031 | 0·034 | 0·37 | 0·03 | 0·04 | 0·44 |
| **Aβ+T+ participants** | Est. | SE | P | Est· | SE | P |
| (Intercept) | 25·41 | 14·10 | *0·071* | 6·96 | 21·07 | 0·74 |
| Time | 8·92 | 3·22 | **0·006** | 11·22 | 4·93 | **0·03** |
| LIBRA | 0·01 | 0·34 | 0·98 | 0·23 | 0·50 | 0·64 |
| Time:LIBRA | 0·005 | 0·064 | 0·94 | 0·05 | 0·08 | 0·57 |

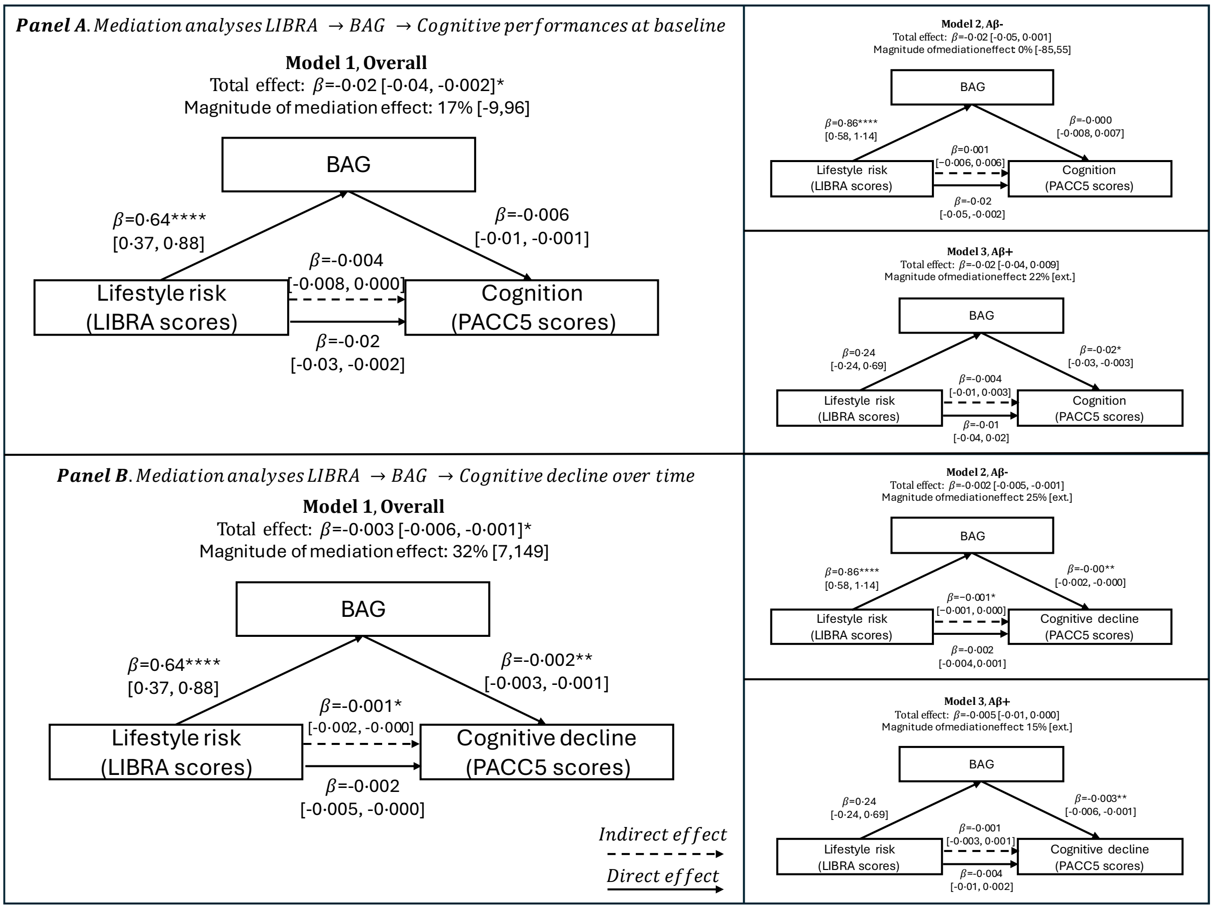

**eFigure 1· Mediation of the association between lifestyle risk (LIBRA) and cognition by brain age gap (BAG) in the CU subsample·**

**eTable 9·** Baseline participants demographics according to clinical groups in ADNI

| | CN | MCI | DAT | Overall | P |
|---|---|---|---|---|---|
| **No· (%)** | 262 (56·8%) | 148 (32·1%) | 51 (11·1%) | 461 (100%) | |
| **Age, y** | 69·3 (66·4, 74·3) | 72·2 (66·8, 77·0) | 75·1 (70·0, 80·2) | 70·9 (66·8, 75·9) | **<0·001** |
| **Female** | 165 (63·0%) | 66 (44·6%) | 18 (35·3%) | 249 (54·0%) | **<0·001** |
| **Education, y** | 17 (16–18) | 16 (14–18) | 16 (14–18) | 16 (15–18) | **0·002** |
| **MMSE score** | 29 (29–30) | 28 (27–29) | 23 (22–24·5) | 29 (27–30) | **<0·001** |
| **Aβ positivity (%)** | 84 (32·1%) | 76 (51·4%) | 41 (80·4%) | 201 (43·6%) | **<0·001** |
| **A/T positivity (%)** | 46 (17·6%) | 65 (43·9%) | 39 (76·5%) | 150 (32·5%) | **<0·001** |
| **FU time, y** | 3·0 (1·2, 4·0) | 2·3 (1·0, 3·3) | 2·0 (0·0, 2·1) | 2·3 (1·0, 4·0) | **<0·001** |
| **FU time MRI, y** | 2·1 (0·0, 4·0) | 2·0 (0·6, 3·0) | 1·0 (0·0, 2·1) | 2·0 (0·0, 3·1) | **<0·001** |

*Baseline values are median (IQR) for continuous data and number of observations (%) for categorical data· Abbreviations: Ab, amyloid; FU, Follow-up, IQR, interquartile; MMSE, mini mental state examination; A/T, amyloid and tau status combined profiles·*

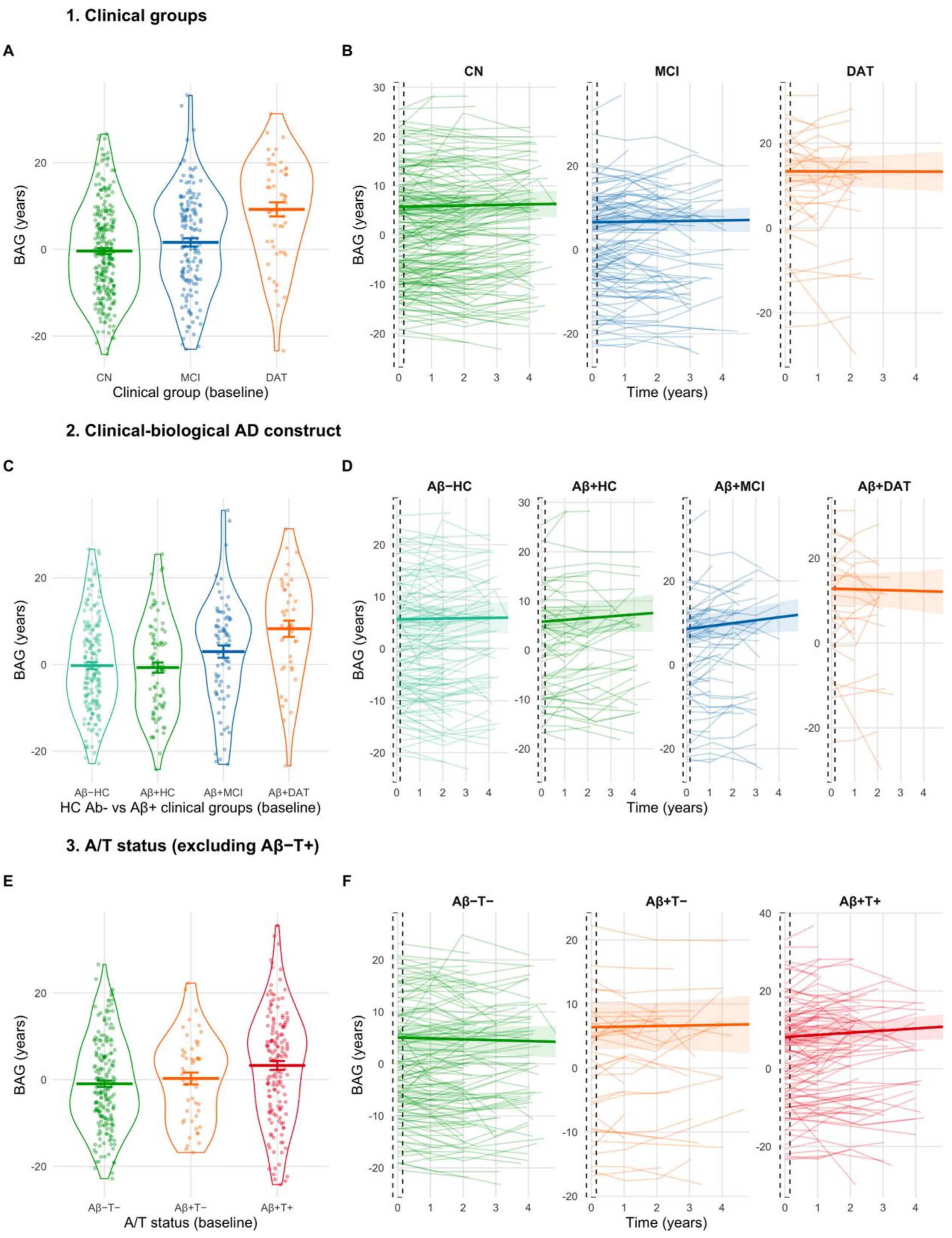

**eFigure 2· Baseline and longitudinal patterns of Brain Age Gap (BAG) across clinical, amyloid, and A/T groups in the ADNI cohort·**

*Notes· Left panels show baseline BAG distribution across clinical groups (A), clinical-biological AD subgroups (C), and A/T profiles (E)· Right panels (B, D, F) display corresponding individual longitudinal BAG trajectories; the dashed box indicates the baseline window (t=0)· Across groups, BAG increased stepwise with clinical severity, was higher in amyloid-positive symptomatic groups, and reached its highest values in Aβ+T1+ individuals· Abbreviations: CN=cognitively normal, SCD=subjective cognitive decline, MCI=mild cognitive impairment, DAT=Alzheimer's type dementia, BAG=brain-age gap, Aβ=β-amyloid, T=tau·*

**eTable 10.** Longitudinal patterns of Brain Age Gap (BAG) across clinical groups in ADNI.

| | Overall | | |
|---|---|---|---|
| **Model 1 - Fixed Effect[a]** | Est· | SE | P |
| (Intercept) | 10·66 | 5·89 | *0·071* |
| Time | 0·67 | 1·10 | 0·544 |
| Group | | | **<0·001** |
| MCI | 0·73 | 1·10 | |
| DAT | 7·46 | 1·67 | |
| Time:Group | | | 0·929 |
| MCI | -0·01 | 0·20 | |
| DAT | -0·15 | 0·38 | |
| **Slopes** | Est· | 95% CI | $P_{adj}$ |
| CN | 0·05 | -0·32, 0·43 | 0·796 |
| MCI | 0·04 | -0·34, 0·42 | 0·796 |
| DAT | -0·09 | -0·69, 0·50 | 0·796 |
| **Pairwise comparisons (BL)** | Est· | 95% CI | $P_{adj}$ |
| CN / MCI | -0·71 | -3·40, 1·97 | 0·52 |
| CN / DAT | -7·29 | -11·39, -3·18 | **<0·001** |
| MCI / DAT | -6·57 | -10·76, -2·38 | **0·001** |
| **Pairwise comparisons (Slopes)** | Est· | 95% CI | $P_{adj}$ |
| CN / MCI | 0·01 | -0·47, 0·49 | 0·957 |
| CN / DAT | 0·15 | -0·77, 1·06 | 0·957 |
| MCI / DAT | 0·13 | -0·79, 1·06 | 0·957 |

**eTable 11.** Longitudinal patterns of Brain Age Gap (BAG) across clinical-biological AD construct (CN Aβ- vs others Aβ+ clinical groups) in ADNI.

| | Overall | | |
|---|---|---|---|
| **Model 1 - Fixed Effect[a]** | Est· | SE | P |
| (Intercept) | 15·78 | 6·90 | **0·023** |
| Time | 0·13 | 1·32 | 0·92 |
| Group | | | **0·001** |
| CN·Aβ+ | 0·11 | 1·40 | |
| MCI·Aβ+ | 2·87 | 1·47 | |
| DAT·Aβ+ | 7·19 | 1·91 | |
| Time:Group | | | *0·06* |
| CN·Aβ+ | 0·30 | 0·24 | |
| MCI·Aβ+ | 0·64 | 0·27 | |
| DAT·Aβ+ | -0·22 | 0·43 | |
| **Slopes** | Est· | 95% CI | $P_{adj}$ |
| CN·Aβ- | -0·18 | -0·60, 0·24 | 0·383 |
| CN·Aβ+ | 0·12 | -0·36, 0·60 | 0·524 |
| MCI·Aβ+ | 0·46 | -0·05, 0·96 | *0·095* |
| DAT·Aβ+ | -0·40 | -1·19, 0·39 | 0·383 |
| **Pairwise comparisons (BL)** | Est· | 95% CI | $P_{adj}$ |
| CN·Aβ- / CN·Aβ+ | -0·48 | -4·24, 3·28 | 0·736 |
| CN·Aβ- / MCI·Aβ+ | -3·64 | -7·60, 0·32 | **0·030** |
| CN·Aβ- / DAT·Aβ+ | -6·93 | -12·11, -1·75 | **0·003** |
| CN·Aβ+ / MCI·Aβ+ | -3·16 | -7·63, 1·30 | *0·09* |
| CN·Aβ+ / DAT·Aβ+ | -6·45 | -11·98, -0·92 | **0·006** |
| MCI·Aβ+ / DAT·Aβ+ | -3·29 | -8·79, 2·21 | 0·14 |
| **Pairwise comparisons (Slopes)** | Est· | 95% CI | $P_{adj}$ |
| CN·Aβ- / CN·Aβ+ | -0·30 | -0·93, 0·33 | 0·318 |
| CN·Aβ- / MCI·Aβ+ | -0·64 | -1·34, 0·07 | 0·107 |
| CN·Aβ- / DAT·Aβ+ | 0·22 | -0·94, 1·37 | 0·614 |
| CN·Aβ+ / MCI·Aβ+ | -0·33 | -1·13, 0·46 | 0·318 |
| CN·Aβ+ / DAT·Aβ+ | 0·52 | -0·68, 1·72 | 0·318 |
| MCI·Aβ+ / DAT·Aβ+ | 0·85 | -0·34, 2·05 | 0·178 |

**eTable 12.** Longitudinal patterns of Brain Age Gap (BAG) across amyloid status (model 1) and A/T status (model 2, excluding Aβ-.T+) in ADNI.

| | Overall | | | CU | | |
|---|---|---|---|---|---|---|
| **Model 1 - Fixed Effect[a]** | Est· | SE | P | Est· | SE | P |
| (Intercept) | 11·96 | 6·01 | **0·047** | 0·80 | 8·16 | 0·922 |
| Time | 1·06 | 1·11 | 0·340 | -0·58 | 1·33 | 0·662 |
| Group [Aβ+] | 2·08 | 1·02 | **0·042** | -0·30 | 1·33 | 0·824 |
| Time:Group [Aβ+] | 0·48 | 0·18 | **0·007** | 0·25 | 0·20 | 0·201 |
| **Model 2 - Fixed Effect[a]** | Est· | SE | P | Est· | SE | P |
| (Intercept) | 11·73 | 6·33 | *0·064* | 3·85 | 8·43 | 0·648 |
| Time | 0·86 | 1·22 | 0·483 | -1·49 | 1·42 | 0·300 |
| Group | | | *0·033* | | | 0·556 |
| Aβ+·T- | 1·35 | 1·67 | | 1·42 | 1·76 | |
| Aβ+·T+ | 3·11 | 1·19 | | -0·84 | 1·68 | |
| Time:Group | | | **0·006** | | | 0·566 |
| Aβ+·T- | 0·25 | 0·29 | | 0·18 | 0·27 | |
| Aβ+·T+ | 0·71 | 0·22 | | 0·24 | 0·25 | |
| **Slopes** | Est· | 95% CI | $P_{adj}$ | Est· | 95% CI | $P_{adj}$ |
| Aβ-·T- | -0·32 | -0·65, 0·01 | **0·031** | -0·14 | -0·47, 0·19 | 0·830 |
| Aβ+·T- | -0·07 | -0·52, 0·38 | 0·713 | 0·04 | -0·40, 0·48 | 0·835 |
| Aβ+·T+ | 0·39 | 0·03, 0·75 | **0·030** | 0·10 | -0·32, 0·52 | 0·829 |
| **Pairwise comparisons (BL)** | Est· | 95% CI | $P_{adj}$ | Est· | 95% CI | $P_{adj}$ |
| Aβ-·T- / Aβ+·T- | -1·65 | -5·74, 2·43 | 0·331 | -1·66 | -5·92, 2·61 | 0·526 |
| Aβ-·T- / Aβ+·T+ | -3·96 | -6·87, -1·04 | **0·004** | 0·52 | -3·54, 4·59 | 0·756 |
| Aβ+·T- / Aβ+·T+ | -2·30 | -6·49, 1·89 | 0·282 | 2·18 | -2·94, 7·30 | 0·526 |
| **Pairwise comparisons (Slopes)** | Est· | 95% CI | $P_{adj}$ | Est· | 95% CI | $P_{adj}$ |
| Aβ-·T- / Aβ+·T- | -0·25 | -0·96, 0·45 | 0·385 | -0·18 | -0·83, 0·47 | 0·763 |
| Aβ-·T- / Aβ+·T+ | -0·71 | -1·24, -0·18 | **0·004** | -0·24 | -0·86, 0·37 | 0·763 |
| Aβ+·T- / Aβ+·T+ | -0·46 | -1·21, -0·29 | 0·209 | -0·07 | -0·86, 0·72 | 0·843 |

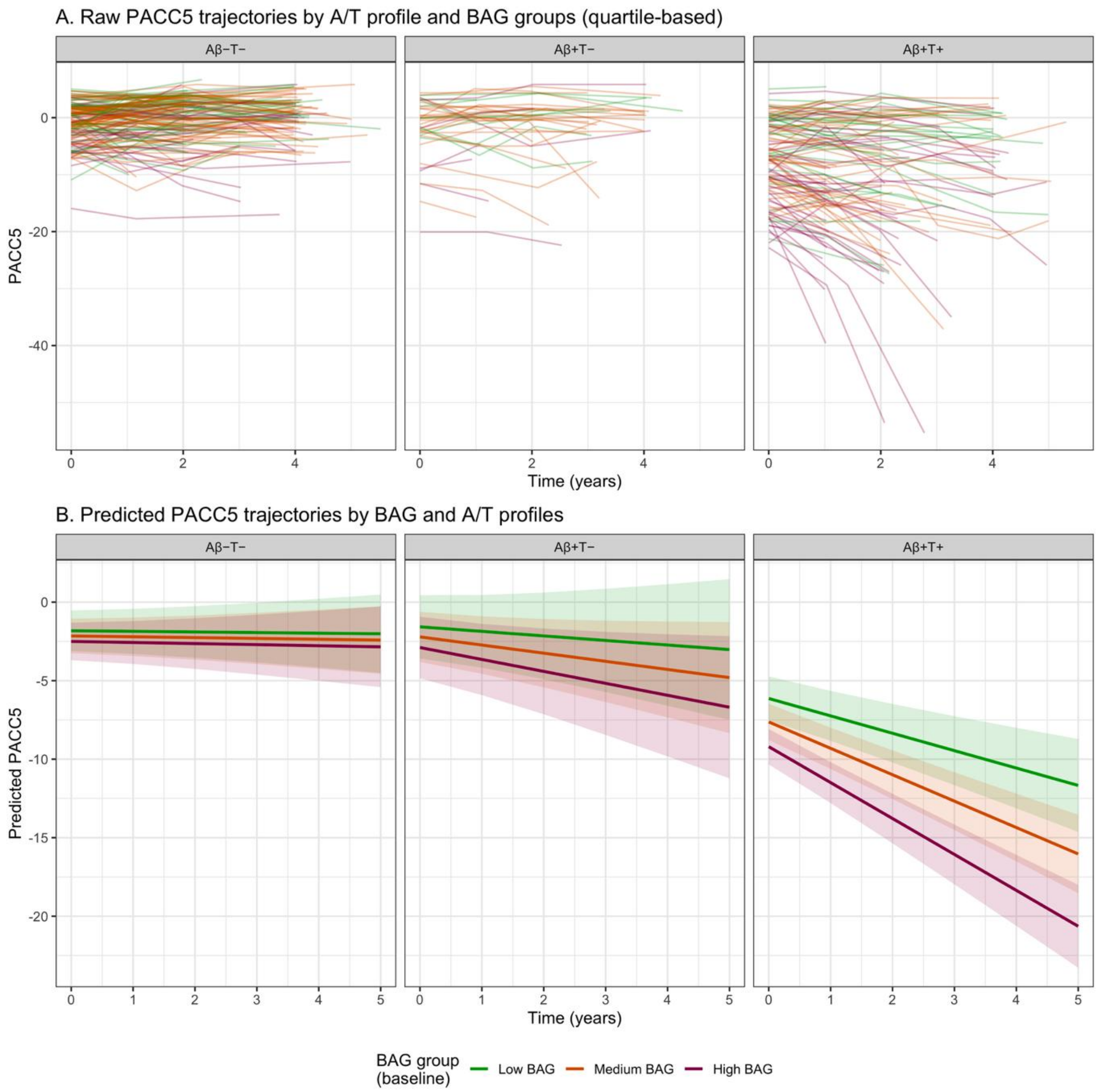

**eFigure 3· Longitudinal PACC5 trajectories according to brain-age gap (BAG) and A/T profile in the ADNI cohort·**

*Notes· Panel A shows observed individual PACC5 trajectories over follow-up, stratified by A/T profile (Aβ−T−, Aβ+T−, Aβ+T+) and baseline brain-age gap (BAG) groups defined by quartiles (low, medium, high)· Panel B shows predicted PACC5 trajectories from linear mixed-effects models including interactions between time, continuous BAG, and A/T profile, adjusted for age, sex, and years of education· Shaded areas represent 95% CIs· Higher BAG indicates an older-appearing brain relative to chronological age· Aβ=β-amyloid· T=tau· BAG=brain-age gap·*

**eTable 13. Longitudinal cognitive performances (PACC5 scores) according to BAG at baseline in ADNI.**

| **Model 1· Overall - Fixed Effect** | Est· | SE | P |
|---|---|---|---|
| (Intercept) | -1·99 | 3·10 | 0·520 |
| Time | 0·42 | 1·19 | 0·725 |
| BAG | -0·14 | 0·02 | **<0·001** |
| Time:BAG | -0·05 | 0·009 | **<0·001** |
| **Model 2· Overall - Fixed Effect** | Est· | SE | P |
| (Intercept) | -2·52 | 2·95 | 0·394 |
| Time | 0·43 | 1·17 | 0·711 |
| BAG | -0·06 | 0·03 | *0·053* |
| BAG:Group [Aβ+] | -0·12 | 0·04 | **0·004** |
| Time:BAG | -0·02 | 0·01 | *0·090* |
| Time:BAG:Group [Aβ+] | -0·05 | 0·02 | **0·002** |
| **Model 3· Overall - Fixed Effect** | Est· | SE | P |
| (Intercept) | -3·76 | 2·97 | 0·206 |
| Time | 1·07 | 1·16 | 0·356 |
| BAG | -0·04 | 0·04 | 0·234 |
| BAG:Group | | | **<0·001** |
| Aβ+T- | -0·04 | 0·08 | |
| Aβ+T+ | -0·14 | 0·05 | |
| Time:BAG | -0·002 | 0·01 | 0·872 |
| Time:BAG:Group | | | **<0·001** |
| Aβ+T- | -0·03 | 0·03 | |
| Aβ+T+ | -0·08 | 0·02 | |
| **Pairwise comparisons (BL)** | Est· | 95% CI | $P_{adj}$ |
| Aβ-·T- / Aβ+·T- | 0·08 | -0·16, 0·32 | 0·439 |
| Aβ-·T- / Aβ+·T+ | 0·25 | 0·11, 0·40 | **<0·001** |
| Aβ+·T- / Aβ+·T+ | 0·17 | -0·06, 0·41 | 0·117 |
| **Pairwise comparisons (Slopes)** | Est· | 95% CI | $P_{adj}$ |
| Low (-10·6 y) | | | |
| Aβ−T− / Aβ+T+ | 0.758 | −0.202, 1.717 | **0.026** |
| Aβ−T− / Aβ+T- | 0.068 | −1.288, 1.425 | 0.951 |
| Aβ+T− / Aβ+T+ | 0.689 | −0.720, 2.099 | 0.193 |
| Mean (+0·8 y) | | | |
| Aβ−T− / Aβ+T+ | 1.618 | 0.935, 2.301 | **<0.001** |
| Aβ−T− / Aβ+T- | 0.397 | −0.533, 1.327 | 0.266 |
| Aβ+T− / Aβ+T+ | 1.221 | 0.248, 2.195 | **<0.001** |
| High (+12·2 y) | | | |
| Aβ−T− / Aβ+T+ | 2.478 | 1.532, 3.425 | **<0.001** |
| Aβ−T− / Aβ+T- | 0.725 | −0.768, 2.218 | 0.193 |
| Aβ+T− / Aβ+T+ | 1.754 | 0.260, 3.247 | **<0.001** |
| Low vs High | | | |
| Aβ−T− | 0.049 | −0.932, 1.030 | 0.951 |
| Aβ+T− | 0.706 | −1.247, 2.658 | 0.326 |
| Aβ+T+ | 1.770 | 0.806, 2.733 | **<0.001** |

**eTable 14**·Risk of clinical progression to incident mild cognitive impairment (iMCI) or major neurocognitive disorder according to BAG·in ADNI

| | | **iMCI_Dem** | | | **iMCI** | |
|---|---|---|---|---|---|---|
| **Model 1** | $N_{obs[event]}$ | HR [95% CI] | P | $N_{obs[event]}$ | HR [95% CI] | P |
| BAG | 460 [36] | 1·05 [1·01, 1·08] | **0·007** | 460 [14] | 1·04 [0·98, 1·09] | 0·20 |
| **Model 1 * Aβ** | $N_{obs[event]}$ | HR [95% CI] | P | $N_{obs[event]}$ | HR [95% CI] | P |
| BAG | 460 [36] | 1·00 [0·93, 1·08] | 0·90 | 460 [14] | 0·98 [0·91, 1·06] | 0·60 |
| BAG * Amyloid status [Aβ+] | 460 [29] | 1·05 [0·97, 1·14] | 0·20 | 460 [8] | 1·09 [0·99, 1·21] | *0·075* |
| **Model 1 * AβT** | $N_{obs[event]}$ | HR [95% CI] | P | $N_{obs[event]}$ | HR [95% CI] | P |
| BAG | 460 [36] | 0·98 [0·91, 1·07] | 0·70 | 460 [14] | 0·98 [0·91, 1·06] | 0·60 |
| BAG * Ab+T- status | 460 [4] | 1·12 [0·98, 1·27] | *0·093* | 460 [4] | 1·14 [0·99, 1·32] | *0·066* |
| BAG * Ab+T+ status | 460 [25] | 1·06 [0·97, 1·16] | 0·20 | 460 [5] | 1·07 [0·96, 1·19] | 0·20 |
